# Long-term effects of a coalmine fire on hospital and ambulance use: an interrupted time series study

**DOI:** 10.1101/2024.05.09.24307097

**Authors:** Tyler J. Lane, Catherine L. Smith, Caroline X. Gao, Jillian F. Ikin, Rongbin Xu, Matthew T.C. Carroll, Emily Nehme, Michael J. Abramson, Yuming Guo

## Abstract

**Background:** In 2014, the Hazelwood coalmine fire in regional Victoria, Australia shrouded nearby communities in smoke for six weeks. Prior investigations identified substantial adverse effects, including increases in the use of health services. In this study, we examined the effects on hospital and ambulance use in the eight years following the fire.

**Methods:** Using Victorian hospital (Jan 2009-Jun 2022) and ambulance (Jan 2013-Dec 2021) data, we conducted an interrupted time series of changes to the rate of hospital admissions, emergency presentations, and ambulance attendances. A categorical exposure model compared two locations, most-exposed Morwell and less-exposed Latrobe Valley, to the rest of regional Victoria. A continuous exposure model used spatial estimates of fire-related PM_2.5_. Analyses were stratified by sex, age group (<65/65+ years), and condition (cardiovascular, respiratory, mental health, injury).

**Results:** There were small but significant increases in overall hospital admissions and emergency presentations across all analyses, but little evidence of change in overall ambulance attendances. Effects varied considerably by condition, with the biggest relative increases observed among hospital admissions for mental health conditions and injuries. While cardiovascular-related hospital admissions and emergency presentations increased post-fire, ambulance attendances decreased.

**Conclusions:** Our findings suggest the Hazelwood coalmine fire likely increased hospital usage. However, it is unclear whether this was due to the direct effects of smoke exposure on health, or the disruptive socioeconomic and behavioural impacts of an environmental disaster that affected how communities engaged with various health services.

## 1 Introduction

In early 2014, embers from a bushfire in regional Victoria, Australia, ignited an opencut coalmine. The mine fire burned for six weeks, shrouding nearby towns in smoke. The most affected region was the Latrobe Valley and Morwell in particular, where residential areas in the southern edge are separated from the coalmine by a highway (see Figure 1). In the fire’s immediate aftermath, residents reported developing symptoms including cough, chest pain, and headache (1). A few months after the fire had been extinguished, the Hazelwood Health Study was established to monitor the ongoing health effects (2).

**Figure 1.**
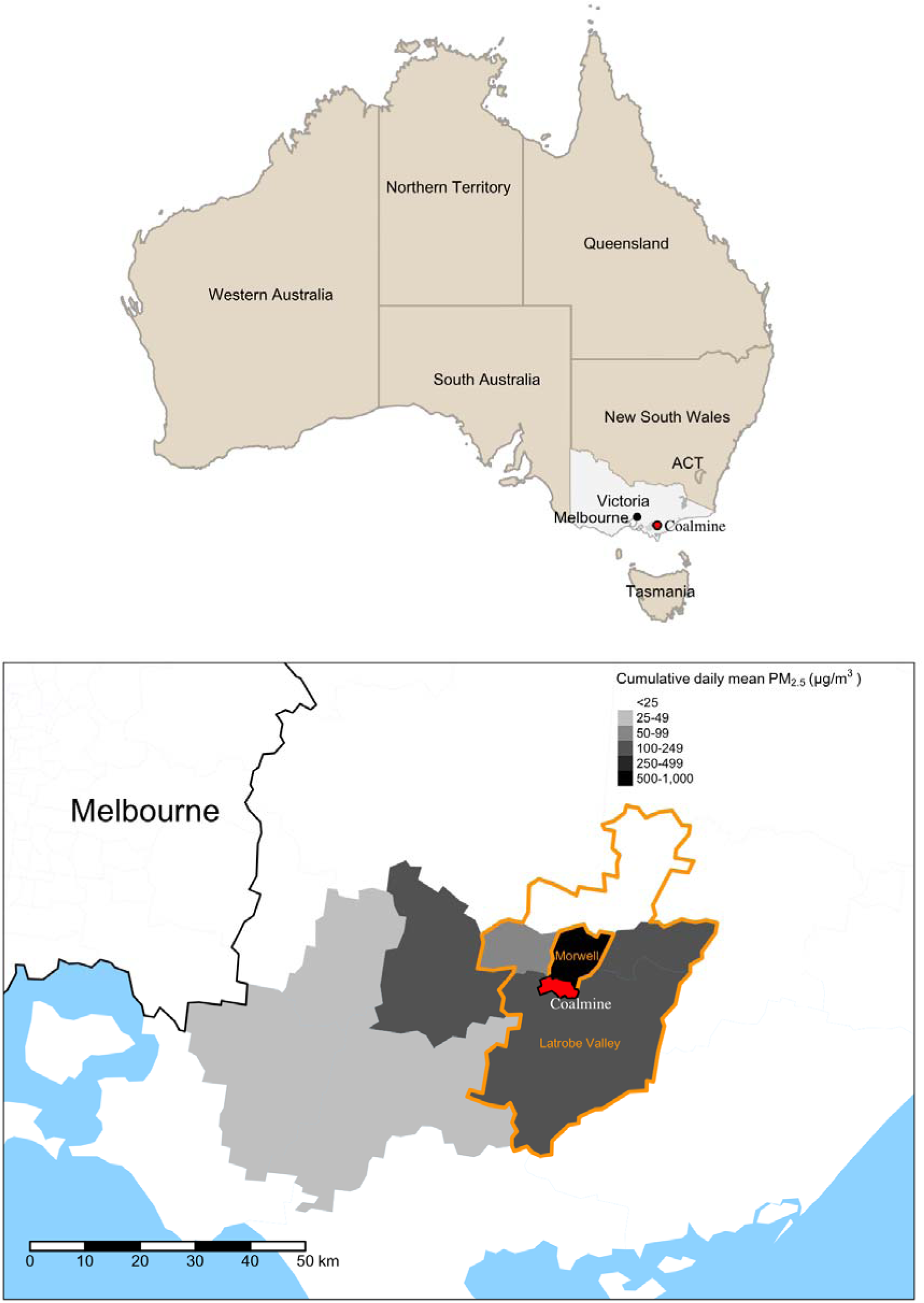
Map of Australia showing the location of the coalmine and a zoom of surrounding areas, with modelled distributions of fire-related PM_2.5_.

Prior to the Hazelwood Health Study, there was limited evidence on health effects of coalmine fire (1,3). Inferences from studies on wildfire smoke and indoor coal use indicated it was likely harmful (3). Coal combustion emits numerous harmful by-products, most notably particulate matter ≤2.5µm (PM_2.5_), which has been linked to health problems in the nervous, respiratory, digestive, cardiovascular, reproductive, and immune systems (4). During the Hazelwood coalmine fire, residential areas of Morwell closest to the mine were estimated to reach a daily mean of 1022µg/m^3^ (5), nearly seven times in excess of Environment Protection Authority Victoria’s 150µg/m^3^ threshold for “extremely poor” air quality (6). There are reasons to believe PM may be more harmful when it originates from coalmine fires (7), including higher concentration of finer particles that can penetrate deeper into the airways, greater composition of oxidative and pro-inflammatory particles, and heavy metals (7,8).

Previous Hazelwood Health Study analyses have linked fire-related PM_2.5_ to increases in GP and respiratory specialist consultations (9), dispensing of medications for respiratory, cardiovascular, and psychiatric conditions (10), hospital admissions and emergency department presentations for all causes as well as respiratory conditions (11), and ambulance attendances for all causes and respiratory conditions (12) both during and shortly after the fire. There is also evidence of medium-term effects, with fire-related smoke exposure being linked to increased emergency presentations for cardiovascular conditions in the following 2.5 years and for respiratory conditions in the following 5 years (13). All-cause hospitalisations were also elevated in the 5 years post-fire, most notably for asthma and chronic obstructive pulmonary disease (14). However, these studies are limited by use of relatively small samples or a focus on short to medium-term outcomes, which were up to five years at most.

In this study, we use more up-to-date population-level hospital and ambulance data to address the following research questions:

1. Did PM_2.5_ exposure from the Hazelwood coalmine fire increase hospital admissions, emergency department presentations, and ambulance attendances in the longer-term (up to eight years post-fire)?
2. Did effects vary by condition?

## 2 Methods

### 2.1 Data

Three data sources were included in this study: the Victorian Admitted Episodes Dataset (VAED) (15), which recorded admissions to Victorian public and private hospitals, rehabilitation centres, extended care facilities, and day procedure centres; the Victorian Emergency Minimum Dataset (VEMD) (16), which recorded emergency department presentations to Victorian public hospitals and others as directed by the Department of Health; and the Ambulance Victoria electronic patient care record data, which recorded ambulance attendances. For simplicity, these are referred to as the hospital, emergency, and ambulance datasets. In all three datasets, each case record included information on admission/presentation/attendance date, diagnosis, sex, age group and postcode.

Hospital and emergency datasets covered January 2009 to June 2022. The ambulance dataset covered January 2013 to December 2021 (pre-2013 were unavailable due to lack of digital data collection in regional areas), excluding a period of industrial action from 25 September 2014 to 31 December 2014. Ambulance data were limited to emergency attendance. While the datasets covered all of Victoria, we excluded Greater Melbourne due to substantial socioeconomic and cultural differences that rendered it largely incomparable to the rest of Victoria. Several regional Statistical Areas were excluded due to small population sizes (Alps – West, French Island, Lake King, Upper Yarra Valley, Wilson’s Promontory).

#### 2.1.1 Exposure groups

We used two separate exposure measures: a categorical exposure and a continuous exposure. The categorical exposure was based on proximity to the mine fire and levels of exposure as indicated by modelled PM_2.5_ data (5), resulting in three groups: 1) Morwell, which experienced the most smoke exposure during the fire (daily mean population-weighted fire-related PM of 32.8 µg/m^3^ during the fire’s peak; see Figures S1 and S2 in the supplementary materials); 2) the rest of the Latrobe Valley (Latrobe Valley hereafter), the local government area in which Morwell sits (daily mean fire-related PM : 3.1µg/m^3^); and 3) rest of regional Victoria, which had little or no exposure to the mine fire smoke.

The continuous exposure model used estimated daily mean fire-related PM_2.5_ at Statistical Area Level 2, a geospatial unit that groups socioeconomically-interactive communities (17). As a result of limited ground-level pollution monitoring, especially during the intensive early stage of the mine fire (1), it was necessary to use estimated PM_2.5_ data, which were developed using retrospective chemical transport modelling that integrated air monitoring, coal combustion, and weather condition data (5).

The locations of Morwell and the Latrobe Valley are illustrated in Figure 1, along with distributions of fire-related PM_2.5_.

#### 2.1.2 Outcomes

There were three outcome groups: hospital admissions, emergency department presentations, and ambulance attendances, summarised as monthly counts (for all and condition-specific service use) by Statistical Area Level 2 geographical boundaries for 2016. Monthly rates were obtained using estimated yearly population counts (18). Condition subgroups were based on ICD-10 codes for primary diagnosis (hospital/emergency) or final code, and consisted of all (A00-Z99), cardiovascular (I***, G45_, G450, G451, G452, G453, G458, G459, G46_, where “*” refers to any character in this position and “_” refers to a space in this position), respiratory (J00*-J99*), mental health (F04-F99), and injuries (S40-S99).

#### 2.1.3 Confounders

Analyses adjusted for mean temperature and humidity (19), number of working days in the week (excluding weekends and public holidays), percentile of Index of Relative Socioeconomic Advantage and Disadvantage (IRSAD) in 2016 (20), long-term trend, and proportion of the population aged 65+ (18).

### 2.2 Statistical analysis

We evaluated the long-term trend of different services use patterns by plotting the monthly times series rate of each outcome for each group.

To assess the Hazelwood coalmine fire’s impact on health service utilization, we used an interrupted time series study design. Time series data were analysed with a quasi-Poisson regression, with event counts as the outcome and logged population as the offset. While interrupted time series often generate estimates on changes to the slope and intercept, we focused solely on the latter to estimate an average exposure effect over an 8-year post-fire period. Categorical and continuous exposure measures were assessed in separate models. Categorical models compared both Morwell and the Latrobe Valley to a counterfactual that was based on secular trends and effects in the rest of regional Victoria. Continuous models compared the effect of fire-related PM_2.5_ to a counterfactual based on secular trends. Analyses were stratified by age (<65/65+ years) and sex (male, female).

All models excluded the months of the mine fire (February-March 2014). Confounders were treated linearly, except for temperature and long-term trend. Since temperature exhibits a nonlinear association with health outcomes, e.g., both low and high temperatures are linked to mortality (21), it was modelled using a natural spline with 2 degrees of freedom. Long-term trends could have been affected by various factors like seasonality, the 2016 introduction of a clinical response model ambulance dispatching system to reduce over-triaging (22), and major disruptions such as the COVID-19 pandemic. To account for these complex trends, we utilised a flexible modelling framework employing a natural spline model with 20 degrees of freedom. Random intercepts were included to allow for variation between Statistical Areas.

Sensitivity analyses were conducted with different parameter settings for natural spline models (i.e., different degrees of freedom), and the inclusion of annual average ambient PM_2.5_ as an additional confounder. The annual average ambient PM_2.5_ exposure for each Statistical Area was generated from satellite-land use regression modelling (23). Data from the year 2020 was excluded from sensitivity analysis models including ambient PM_2.5_ because of the countervailing effects of the 2019-2020 Black Summer bushfire season – during which period widespread smoke exposure likely increased medical service use – and the COVID-19 pandemic – when lockdowns drastically reduced health service use (24–26). Analyses were conducted in R (27) using RStudio (28).

### 2.3 Ethics

This study was approved by Monash University Human Research Ethics Committee (MUHREC) as part of the Hazelwood Adult Survey & Health Record Linkage (approval number: 25680).

## 3 Results

### 3.1 Descriptives

Time series plots for overall hospital admissions, emergency presentations, and ambulance attendances are provided in Figure 2; for demographic and condition subgroups, see Figures S3-S7 in the Supplementary Materials. Table 1 presents the mean and standard deviation for monthly service use rates pre- and post-fire. Morwell had higher utilisation rates for all three health services, followed by Latrobe Valley. Trends and seasonal patterns were similar across all three groups. All three services saw reductions in 2020, corresponding to the COVID-19 pandemic and lockdowns.

**Figure 2.**
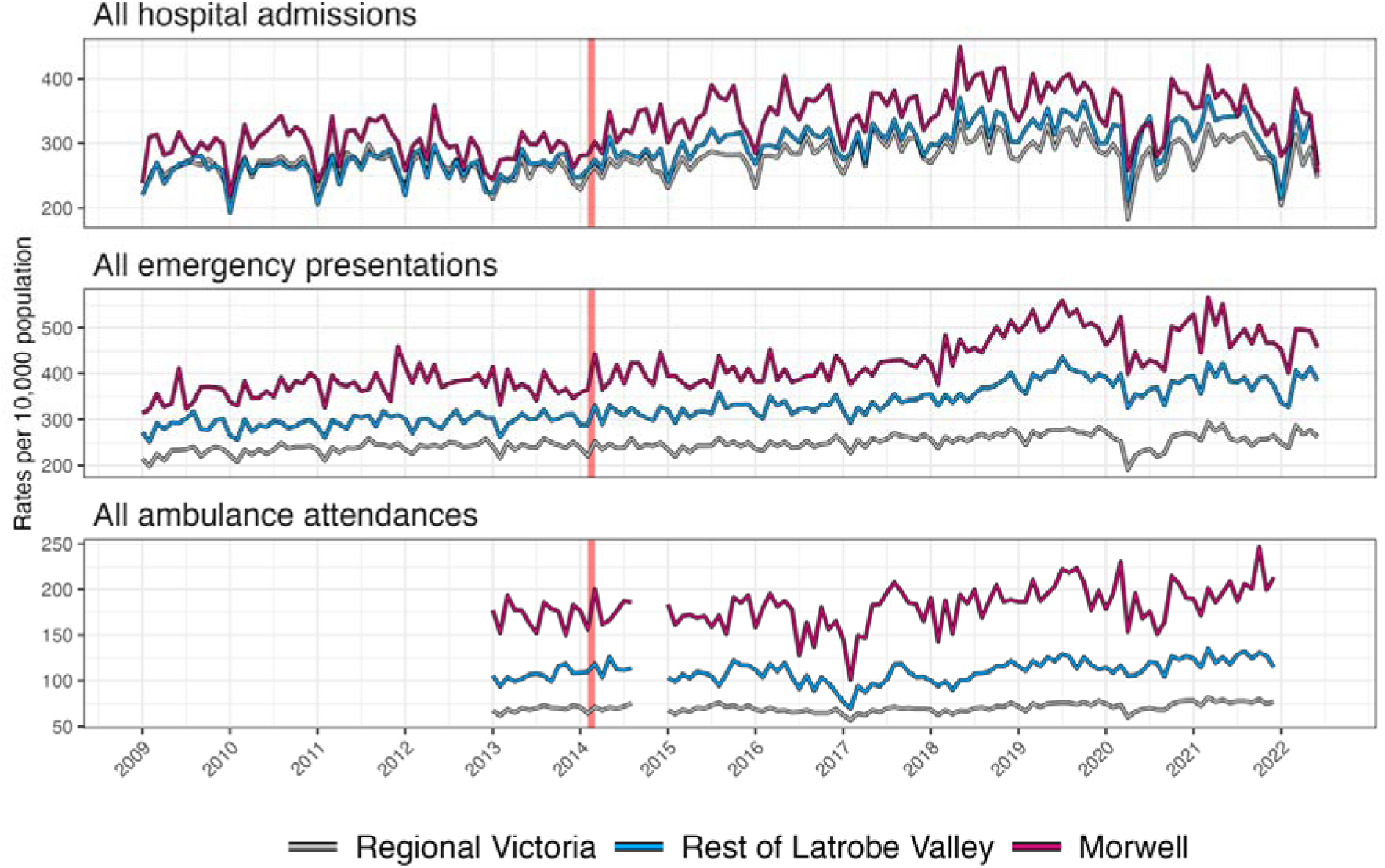
Trends of each service use over time.

**Table 1.**
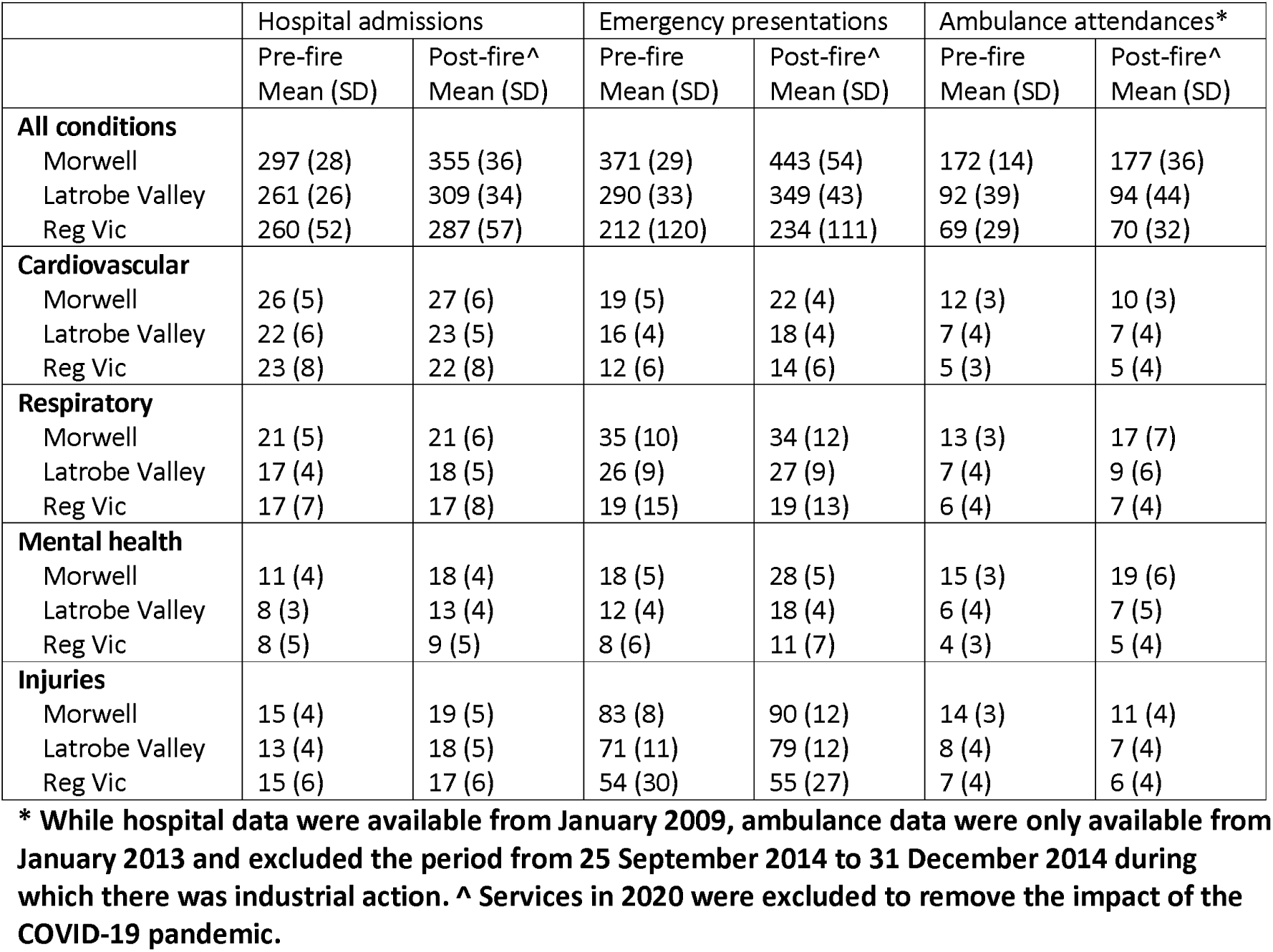
Distribution of monthly service use per 10,000 residents before and after the Hazelwood coalmine fire by exposure group across SA2 areas.

|  | Hospital admissions |  | Emergency presentations |  | Ambulance attendances* |  |
| --- | --- | --- | --- | --- | --- | --- |
|  | Pre-fire<br>Mean (SD) | Post-fire^<br>Mean (SD) | Pre-fire<br>Mean (SD) | Post-fire^<br>Mean (SD) | Pre-fire<br>Mean (SD) | Post-fire^<br>Mean (SD) |
| **All conditions** |  |  |  |  |  |  |
| Morwell | 297 (28) | 355 (36) | 371 (29) | 443 (54) | 172 (14) | 177 (36) |
| Latrobe Valley | 261 (26) | 309 (34) | 290 (33) | 349 (43) | 92 (39) | 94 (44) |
| Reg Vic | 260 (52) | 287 (57) | 212 (120) | 234 (111) | 69 (29) | 70 (32) |
| **Cardiovascular** |  |  |  |  |  |  |
| Morwell | 26 (5) | 27 (6) | 19 (5) | 22 (4) | 12 (3) | 10 (3) |
| Latrobe Valley | 22 (6) | 23 (5) | 16 (4) | 18 (4) | 7 (4) | 7 (4) |
| Reg Vic | 23 (8) | 22 (8) | 12 (6) | 14 (6) | 5 (3) | 5 (4) |
| **Respiratory** |  |  |  |  |  |  |
| Morwell | 21 (5) | 21 (6) | 35 (10) | 34 (12) | 13 (3) | 17 (7) |
| Latrobe Valley | 17 (4) | 18 (5) | 26 (9) | 27 (9) | 7 (4) | 9 (6) |
| Reg Vic | 17 (7) | 17 (8) | 19 (15) | 19 (13) | 6 (4) | 7 (4) |
| **Mental health** |  |  |  |  |  |  |
| Morwell | 11 (4) | 18 (4) | 18 (5) | 28 (5) | 15 (3) | 19 (6) |
| Latrobe Valley | 8 (3) | 13 (4) | 12 (4) | 18 (4) | 6 (4) | 7 (5) |
| Reg Vic | 8 (5) | 9 (5) | 8 (6) | 11 (7) | 4 (3) | 5 (4) |
| **Injuries** |  |  |  |  |  |  |
| Morwell | 15 (4) | 19 (5) | 83 (8) | 90 (12) | 14 (3) | 11 (4) |
| Latrobe Valley | 13 (4) | 18 (5) | 71 (11) | 79 (12) | 8 (4) | 7 (4) |
| Reg Vic | 15 (6) | 17 (6) | 54 (30) | 55 (27) | 7 (4) | 6 (4) |

\* While hospital data were available from January 2009, ambulance data were only available from January 2013 and excluded the period from 25 September 2014 to 31 December 2014 during which there was industrial action. ^ Services in 2020 were excluded to remove the impact of the COVID-19 pandemic.

### 3.2 Hazelwood coalmine fire effects on healthcare use

The results for individual interrupted time series models are summarised in Figure 3 and Tables S1-S3. The addition of background PM_2.5_ to models in the sensitivity analyses did not meaningfully affect the relationship between residential area fire-related smoke exposure and health service use. These results can be found in the supplementary materials (Figures S8-S9 and Tables S4-S9).

**Figure 3.**
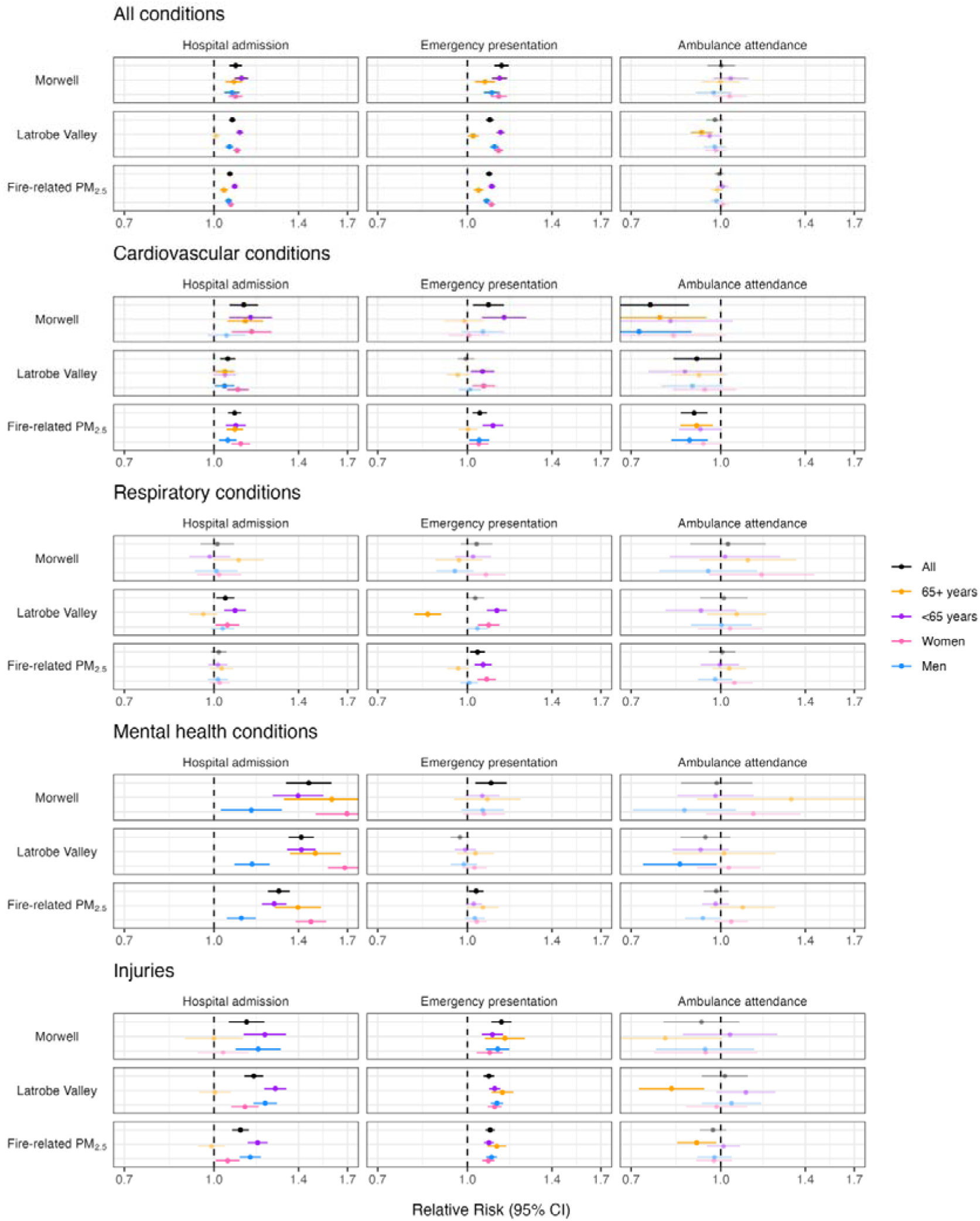
Changes in hospital admission, emergency presentations, and ambulance attendance in the eight years following the Hazelwood coalmine fire; fire-related PM_2.5_ (10µg/m^3^) was evaluated in separate models from those comparing Morwell and (rest of) Latrobe Valley to the rest of regional Victoria.

#### 3.2.1 All conditions

In the 8 years following the Hazelwood coalmine fire, all smoke exposure measures were associated with increased hospital admissions and emergency presentations. Compared with pre-fire trends and relative to changes in the rest of regional Victoria, the rate of hospitalisation increased 9% (95%CI: 1.06-1.12) in Morwell and 8% (95%CI: 1.06-1.09) in the Latrobe Valley, while the rate of emergency presentations increased 15% (95%CI: 1.11-1.18) in Morwell and 9% (1.08-1.11) in the Latrobe Valley. There were significant increases within all subgroups except hospital admissions among older persons in the Latrobe Valley. Though overall effects were generally small (RR: ≤1.15), they were larger among women and younger persons. There were no detectable effects among ambulance attendances except for a reduction among older persons in the Latrobe Valley.

#### 3.2.2 Cardiovascular conditions

Cardiovascular-related hospital admissions increased for nearly every group in each analysis. Effects were larger in most-exposed Morwell than less-exposed Latrobe Valley, which aligns with an exposure dose-related increase. The time series plots in Figures S3-S7 indicate that these peaked in early 2018, particularly among younger persons and men.

There was less consistency among emergency presentations, where we observed overall increases in both Morwell and the fire-related PM_2.5_ model. While there was not a detectable effect on emergency presentations overall in the Latrobe Valley, there were increases among younger persons and women. Conversely, cardiovascular-related ambulance attendances decreased across all models and subgroups, though this was not always statistically significant.

#### 3.2.3 Respiratory conditions

There was little consistency in respiratory-related health service use effects. We only observed effects among hospital admissions in the Latrobe Valley, where there were increases overall and among younger persons and women. For emergency presentations, overall point estimates were positive for each exposure measure, though only significant in the fire-related PM_2.5_ model. There were subgroup increases in younger persons and women in both the Latrobe Valley and the fire-related PM_2.5_ model. There was also a substantial reduction in respiratory-related emergency presentations among older persons in the Latrobe Valley. There were also no detectable effects on ambulance attendances.

Time series plots (Figures S3-S7) indicate that of all conditions, health service use for respiratory conditions exhibited the clearest seasonality, with peaks in winter months (June-August) and troughs in summer (December-February). Notably, while summertime incidence of both hospital admissions and emergency presentations were similar across Morwell, the Latrobe Valley, and the rest of regional Victoria, the winter peaks were much higher for respiratory-related hospitalisations in Morwell and emergency presentations in Morwell and Latrobe Valley, both before and after the mine fire.

#### 3.2.4 Mental health conditions

Mental health-related hospital admissions increased for all groups across all exposure measures, and represented the largest relative risk increases across all analyses and all groups, with the biggest effects among older persons and women. Time series plots (Figures S3-S7) indicate that in Morwell, the increase was most prominent in the three years following the mine fire, as well as an overall peak in late 2018 that was also observable among younger persons and men. There was also a notable peak in mental health-related hospital admissions among older persons in late 2019.

There were overall increases in mental health-related emergency presentations in Morwell and in the PM_2.5_ model. While there were no detectable effects within subgroups, point estimates were generally positive in Morwell and in the fire-related PM_2.5_ model.

There were no detectable increases in mental health-related ambulance attendances, though point estimates for older persons indicated a substantial increase. There were reductions in ambulance attendances for men in the Latrobe Valley; while non-significant, the point estimate in Morwell was similar in magnitude.

#### 3.2.5 Injuries

With a few exceptions, injury-related hospital admissions and emergency presentations increased post-fire across exposure measures and subgroups; the exceptions were no detectable change in hospital admissions among older persons in any exposure group or among women in Morwell. Effect sizes were similar across exposure measures for emergency presentations, but the overall effect was slightly larger in Morwell. Time series plots in Figures S3-S7 indicate the clearest divergence was in emergency presentations, which starting around mid-2017 turned upward in Morwell and Latrobe Valley but downward in the rest of regional Victoria, which was observed across all groups. There were few detectable changes in injury-related ambulance attendances aside from decreases among older persons, which were significant in all groups except Morwell.

## 4 Discussion

### 4.1 Mine fire effect on hospital, emergency department, and ambulance use

Our findings suggest that the Hazelwood coalmine fire increased hospital admissions and emergency presentations in the following 8 years, though by a relatively small amount. What is not clear is whether these were direct effects of smoke exposure on health, or indirect socioeconomic effects of the fire, such as the closure of the mine and coal-fired powerplant in 2017 or behavioural changes in how individuals engaged with health systems.

If the increase in medical service use was due to direct effects of the fire on health, we would have expected stronger evidence of a dose-exposure response, with larger effects in the most-exposed site (Morwell) relative to the second most-exposed site (rest of the Latrobe Valley). Yet aside from hospital admissions and emergency presentations for cardiovascular conditions, effect magnitudes were similar across both sites and sometimes larger in the Latrobe Valley. There are two possible explanations. The first is that, independent of its health effects, the mine fire led to behavioural changes in how people engaged with healthcare services, possibly by increasing social acceptance of using them or greater salience about their availability. For instance, a previous Hazelwood Health Study analysis found asthma diagnoses increased among men but not women. While men may have had more smoke exposure due to gendered division of labour that increased time outdoors, the increase in asthma diagnoses could have been a result of men seeking medical advice for underlying asthma rather than a direct consequence of smoke exposure (29). Further, the Latrobe Valley Health Assembly, which was established in 2016 partly in response to the mine fire, has actively promoted Hazelwood Health Study findings about the consequences of smoke exposure. This may have prompted some individuals to seek medical care, regardless of whether their health concern was a result of the fire. As the Latrobe Health Assembly serves the larger Latrobe Valley, behavioural changes in how people access healthcare services may have been similar across sites. One example is the Assembly’s *Mental Health Awareness Program*, which was dedicated towards improving community awareness of mental health issues (30); perhaps relatedly, we found the mine fire had similar effects on mental health-related hospital admissions in both Morwell and the Latrobe Valley.

Alternatively, smoke from the mine fire may have worsened health in proportion to exposure, but high levels of socioeconomic deprivation in Morwell may have suppressed any increase in health service use. It is important to note that health service use is not a direct measure of health. For many conditions such as the common cold or minor injuries, most people do not require treatment, much less need to go to hospital or call an ambulance. We can therefore think of some healthcare use as discretionary, regardless of whether it is considered unnecessary or beneficial. However, condition severity and urgency are not the only factors influencing the decision to seek healthcare. Cost, accessibility, cultural factors such as trust, and socioeconomic circumstances all play a role (31). People with financial constraints will have reduced capacity to increase health service use, even if they were made unwell by the mine fire. For instance, during the COVID-19 lockdown period (1 April to 15 October 2020), new cancer notifications in Victoria decreased by 5.3% in the most socioeconomically disadvantaged areas and by 14.7% in the least disadvantaged areas (32); in other words, more disadvantaged areas appeared to have less discretionary health service use to give up during the lockdowns. Further, the largest reductions were among prostate cancer and melanoma (32), which are typically detected early when symptoms are not yet serious and there is more discretion about whether to seek medical advice, while there were no detectable changes in lung cancer, which is typically diagnosed at end stages when there is less discretion about whether to seek medical advice. It is important to reiterate that the effects on new cancer notifications were not due to a change in underlying cancer incidence, but influence on the decision to seek medical attention for cancer-related symptoms. In the case of Morwell, it is one of the most disadvantaged communities in Australia, ranked in the 4^th^ percentile nationally and 2^nd^ percentile in Victoria for socioeconomic deprivation in 2011 (33). In 2021, it was in the 2^nd^ percentile nationally and was the second most deprived area in Victoria (34). Morwell is also considerably more disadvantaged than the rest of the Latrobe Valley, aside from Moe/Newborough, as illustrated in Figure S4. Therefore, while Morwell experienced the most smoke exposure, healthcare service use may have been suppressed due to socioeconomic constraints that limited any increase in discretionary health service use, even if where health worsened.

The Hazelwood coalmine fire’s effects on ambulance attendances provide additional evidence that much of the change in health service use could be due to socioeconomic and behavioural effects. Despite increases in hospital admissions and emergency presentations, there were no detectable increases in ambulance attendances. And among cardiovascular conditions, ambulance attendances decreased across all groups while hospital admissions and emergency presentations increased. This suggests change in how people sought services. The cost of ambulances may be one factor. Victoria has a subscription model for ambulance coverage; those without a subscription or government concession must pay for each call-out. In regional Victoria, 2023/2024 ambulance call-out fees were $586 for treatment without transport and $2,004 for emergency road transport (35). Socioeconomic changes in Morwell may have dissuaded individuals from maintaining their subscriptions, while also putting others within the concession card category, i.e., reaching pension-age or receiving social security. It is worth noting that despite its socioeconomic challenges, health service use including ambulance attendance was highest in Morwell (see Figures 2 and S3-S7 and Table 1). This could be the result of poorer general health in Morwell, increasing both the underlying need for non-discretionary healthcare as well as the financial constraints to accessing additional healthcare.

### 4.2 Effects by condition type

We hypothesised that if smoke from the mine fire worsened health and increased health service use, we would observe a larger effect in Morwell, the highest exposure site, than in Latrobe Valley, the less-exposed site. Yet this was only observed among cardiovascular conditions. However, as noted above, the patterns within cardiovascular effects were peculiar: while both hospital admission and emergency department admissions increased most prominently in Morwell, it also had the largest *decrease* in ambulance attendances. This may reflect a real increase in cardiovascular conditions, but it may also be the result of a behaviour change in which health services were accessed for cardiovascular conditions after the fire, e.g., driving to hospital rather than calling an ambulance. Yet it remains unclear why we only observed such contrasting and potentially off-setting effects between health services among cardiovascular conditions. Further, these results, which are the average estimated effect in the 7.5 to 8 years post-fire, contrast with earlier Hazelwood Health Study findings, which suggested effects on cardiovascular health were short-lived (13,14). However, previous studies were limited to the first five years post-fire, or used linked cohort data rather, which, while allowing for the adjustment of individual-level confounders, limited statistical power. It is also possible that smoke exposure had a two-pronged effect: an immediate increase in cardiovascular-related mortality (36) resulting in a “depletion of the susceptibles” (37), whereby the surviving population is less prone to cardiovascular harms, combined with a latent increase in cardiovascular problems, or an unrelated event that exacerbated smoke exposure effects several years after the fire. Time series plots indicated several peaks in cardiovascular-related hospital admissions starting around 2018 in Morwell. This would have been outside the data collection period for previous Hazelwood Health Study analyses that included analysis of cardiovascular effects. Notably, this peak follows the 2017 closures of the coalmine and the coal-fired powerplant in Morwell. It is also worth noting a similar peak in mental health-related hospital admissions in late-2018, just a few months after the early-2018 peak in cardiovascular-related hospital admissions. Both may be interrelated consequences of this economic change and major social upheaval. Previous Hazelwood Health Study analyses found strong relationships between mental distress and somatic symptoms, including some that are related to cardiovascular conditions (pain in limbs, chest pain, dizziness, fainting spells, palpitations) (38). Alternatively, the initial and Omicron COVID-19 waves (April 2020 and January 2022) were followed by sharp reductions in heart health checks (39). While this would not necessarily varied across regional Victoria, it may have left those exposed to mine fire smoke more vulnerable to underlying cardiovascular issues that would arise without check-up. However, this would still not explain why cardiovascular-related hospital admissions and emergency presentations increased while ambulance attendances decreased.

Another difference between the present findings and previous Hazelwood Health Study analyses was we found few effects on healthcare use for respiratory conditions. The one exception was the fire-related PM_2.5_ model, in which we detected increases in emergency presentations overall and among women and younger persons. Previous Hazelwood Health Study analyses consistently found long-term increases in respiratory symptoms (29,40,41), as well as medium-term increases in emergency department presentations for respiratory conditions (13,14) and poorer lung function in relation to smoke exposure (42). However, recent analysis suggests that the effects on lung function have attenuated over time (43), possibly indicating recovery. It is also worth noting that in this analysis, we averaged effects in the eight years post-fire; attenuation due to recovery could mask short and medium-term effects.

The largest effects in this analysis were observed among hospital admissions for mental health conditions. There were also smaller overall increases in mental health-related emergency presentations in Morwell and in the fire-related PM_2.5_ model. Previous Hazelwood Health Study analyses found substantial increases in hospital admissions, emergency presentations, and ambulance attendances for mental health conditions during the mine fire (44), and that smoke exposure was associated with elevated psychological distress 5-6 years post-fire (45). But as with other conditions, it remains possible that experiences with hospital services during the mine fire affected later behaviour, making it more acceptable to seek these out for mental health issues. Notably, in this study we found the effect on hospital admissions for mental health conditions were considerably larger for women than men. However, previous Hazelwood analyses found men were more vulnerable to delayed-onset psychological distress, but were otherwise similar to women on levels of recovery and chronic trajectories of distress (46). This further suggests an interaction between mental health effects of the mine fire and the changing socioeconomic and demographic context. While mental health-related hospitalisations increased shortly after the mine fire, it is also worth noting a late-2018 peak (see Figures S3-S7). As noted in the discussion about cardiovascular-related effects, the increase in mental health-related health service use may be related to the closure of the coalmine and coal-fired power plant.

Further, while the post-mine fire increases in mental health hospitalisations were the largest effects in the study, they still only accounted for a fraction of total hospitalisations (see Table 1).

The increases in hospital admissions and emergency presentations for injuries were unexpected, though there was some precedent. During the mine fire, there was an increase in deaths due to injury (36). There is a growing body of evidence suggests ambient air pollution and smoke from fire events impair cognitive functioning, which could affect behaviour and judgement (47–50), which may increase the risk of injury (36). Areas with the most smoke exposure from the Hazelwood coalmine fire also had substantial and sustained reductions in children’s standardised numeracy and literacy test scores, though it was unclear whether this was due to smoke effects on cognitive development or the lingering effects of school disruption and closures during the fire (51). The neurological effects of the Hazelwood coalmine fire should become a topic for future investigation.

### 4.3 Strengths and limitations

This study had numerous strengths. We used administrative healthcare data from three different sources covering all of regional Victoria, which was effectively population-level data and offered sufficient statistical power for subgroup analyses by condition type, age group, and sex. The effects of the coalmine fire were evaluated using both categorical/ordinal and continuous measures. Analyses adjusted for important confounders including weather, work days, and baseline socioeconomic and demographic factors.

This study also had several limitations. Hospital, emergency department, and ambulance service use are not direct markers of health or healthcare needs. Ambulance data had reduced coverage than hospital and emergency data, providing only one year pre-mine fire, plus the loss of several months of data due to industrial action shortly after the fire. While our analytical approach, the interrupted time series, is considered one of the most robust quasi-experimental designs for causal inference (52), i.e., being able to attribute an *effect* (changes in health service use) to a *cause* (the Hazelwood coalmine fire), it is nevertheless limited for determining underlying mechanisms, such as whether the changes were due to health effects of smoke exposure or the broader socioeconomic effects. Adjustments for socioeconomic confounders only accounted for underlying differences between areas, not whether they moderated the mine fire’s effects as in the suppression of healthcare-seeking behaviour due to socioeconomic deprivation.

## 5 Conclusions

Our findings suggested the Hazelwood coalmine fire was associated with increased healthcare service use in the longer term. However, some of the effects did not align with previous Hazelwood Health Study findings, such as stronger evidence for an effect on cardiovascular conditions than respiratory conditions. As healthcare service use is not a direct measure of health, it remains unclear how much of the increases were due to direct health effects of smoke exposure compared to indirect effects on how the community engaged with healthcare services in the aftermath.

## Data Availability

As the data contain personal identifiers and as per our agreements with data custodians, they are not publicly available.

## SUPPLEMENTARY MATERIALS

### 1 Fire-related PM_2.5_

In the main document’s Introduction, we compared daily mean fire-related PM_2.5_ in Morwell (32.8 µg/m^3^ ) and the rest of the Latrobe Valley (3.1 µg/m^3^ ) using the “peak fire period”, which we define as 9 February 2014 to 6 March 2014. Using modelled PM_2.5_ data from Luhar et al. 2020 (5), this has been illustrated in Figure S1 (53). The area shaded pink highlights the timeframe in which there were clearly visible peaks in fire-related PM_2.5_. Yet from 7 March 2014 onward, model suggests very low amounts of fire-related PM_2.5_.

**Figure S1.**
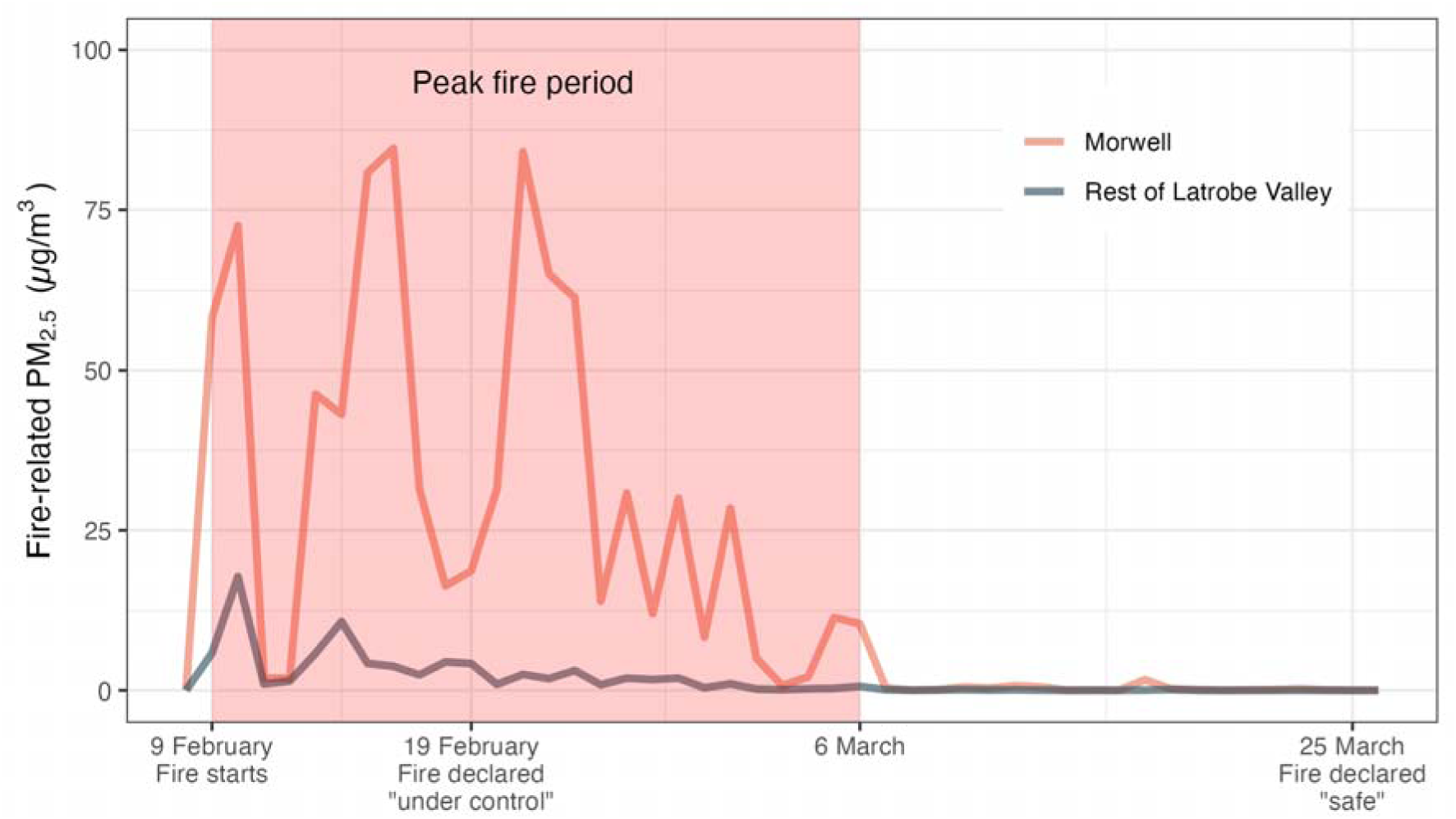
Daily mean fire-related PM_2.5_ during the Hazelwood coalmine fire in Morwell and the rest of the Latrobe Valley.

Figure S2 builds on the map in Figure 1 and the time series in Figure S1 to illustrate how much greater smoke exposure from the Hazelwood coalmine fire was in Morwell and – to a lesser extent – the Latrobe Valley. This figure uses a longer time frame for which fire-related PM_2.5_ was estimated (2 February 2014 to 28 March 2014). Each line underneath the distribution curve stands in for a Statistical Area at Level 2. Morwell, which is red, is far to the right, indicating extreme amounts of cumulative daily mean fire-related PM_2.5_. The rest of the Latrobe Valley, in yellow, follows, though distantly.

**Figure S2.**
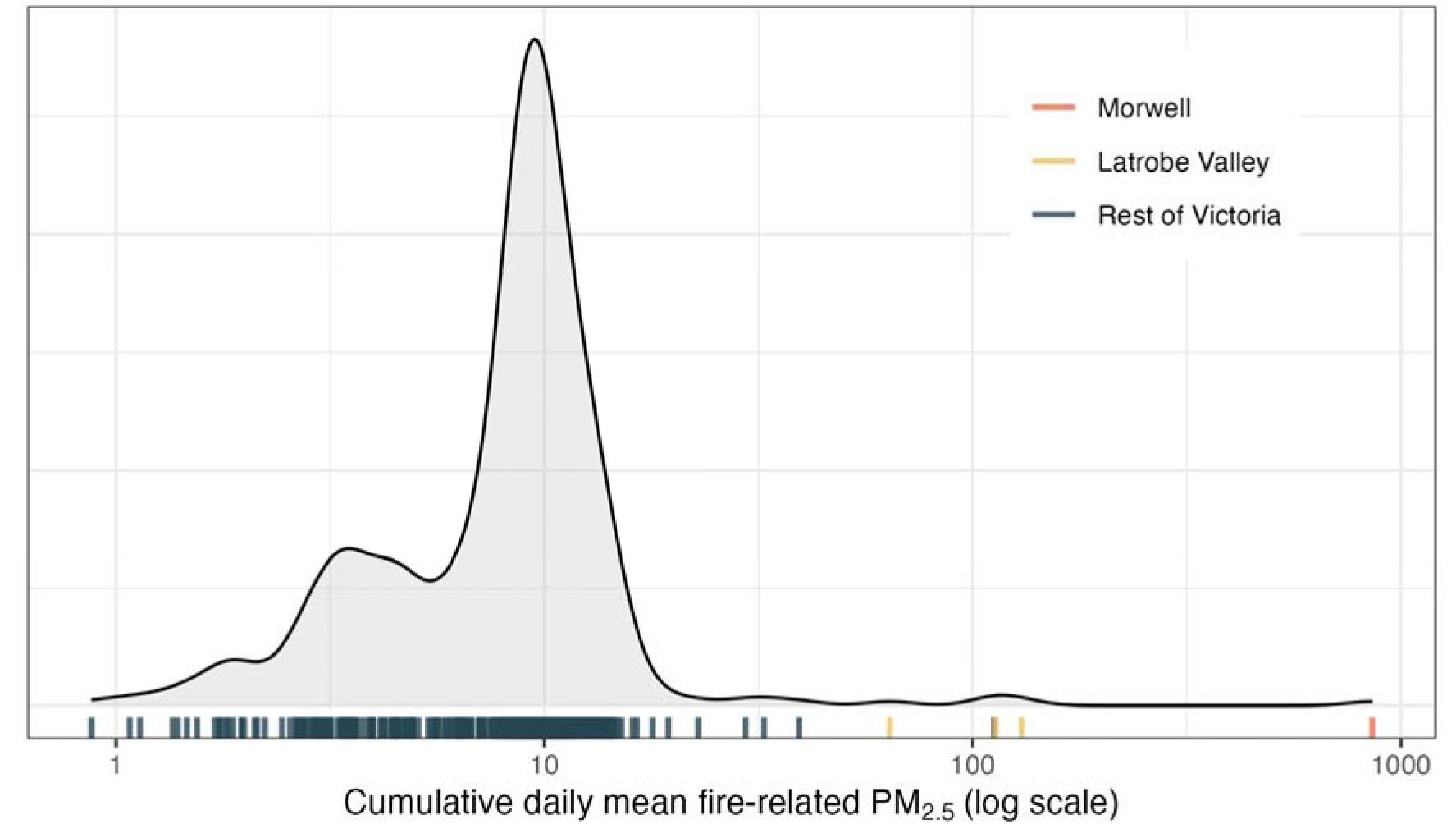
Distributions of cumulative daily mean coalmine fire-related PM_2.5_ by Statistical Area at Level 2.

### 2 Time series plots

**Figure S3.**
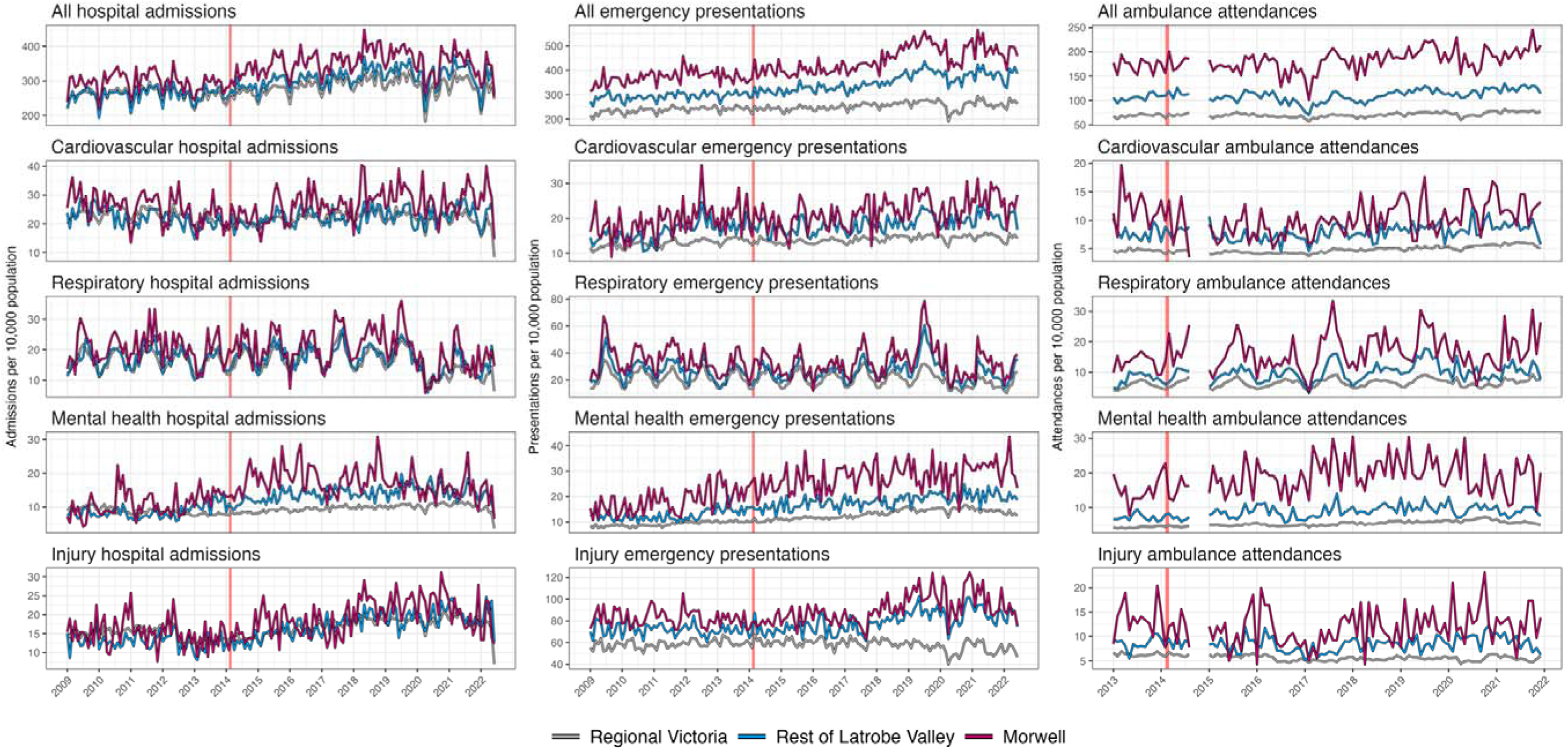
Crude time series plots indicating total monthly hospital admissions, emergency department presentations, and ambulance attendances, by condition, TOTAL.

**Figure S4.**
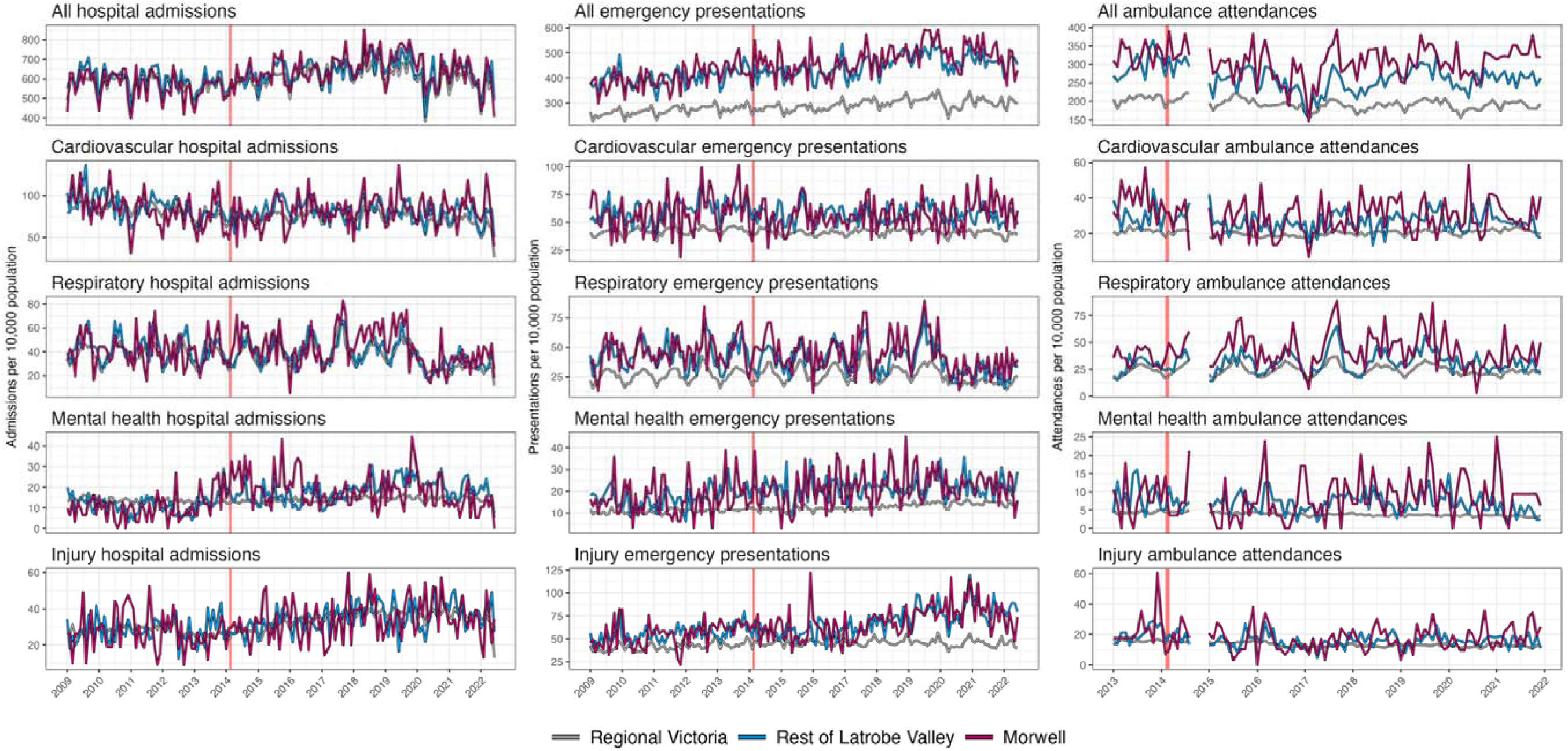
Crude time series plots indicating total monthly hospital admissions, emergency department presentations, and ambulance attendances, by condition, AMONG THOSE AGED 65+ YEARS.

**Figure S5.**
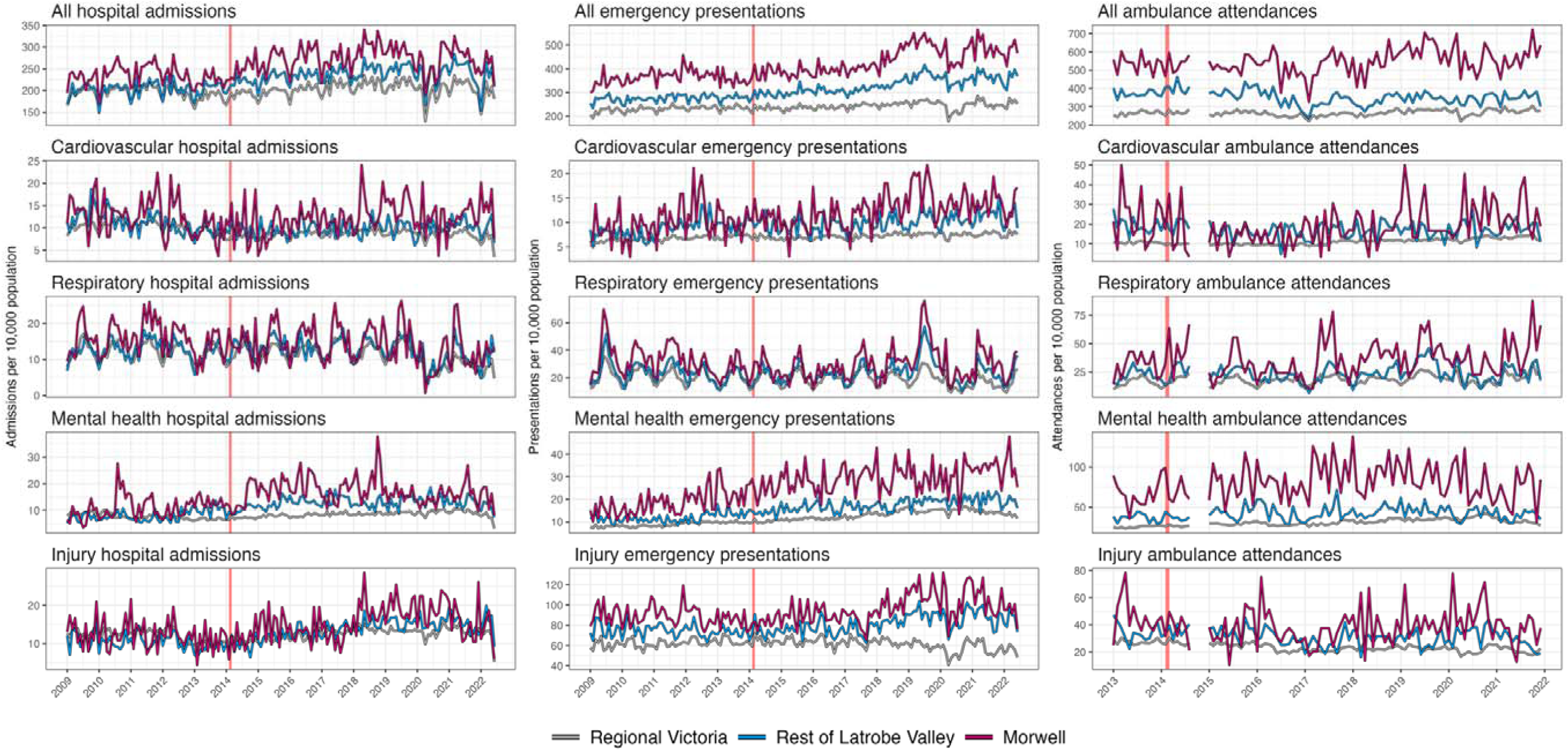
Crude time series plots indicating total monthly hospital admissions, emergency department presentations, and ambulance attendances, by condition, AMONG THOSE AGED <65 YEARS.

**Figure S6.**
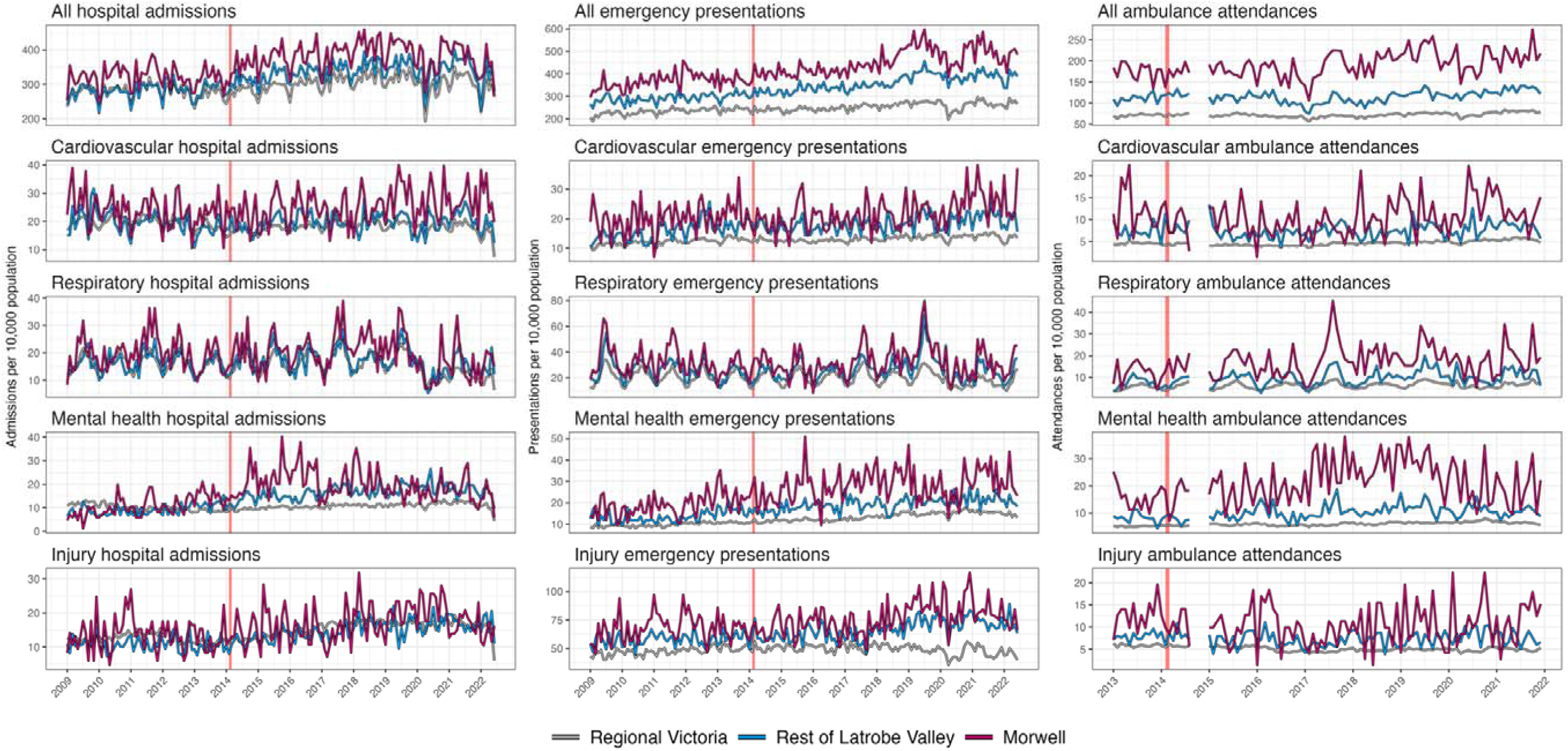
Crude time series plots indicating total monthly hospital admissions, emergency department presentations, and ambulance attendances, by condition, AMONG WOMEN.

**Figure S7.**
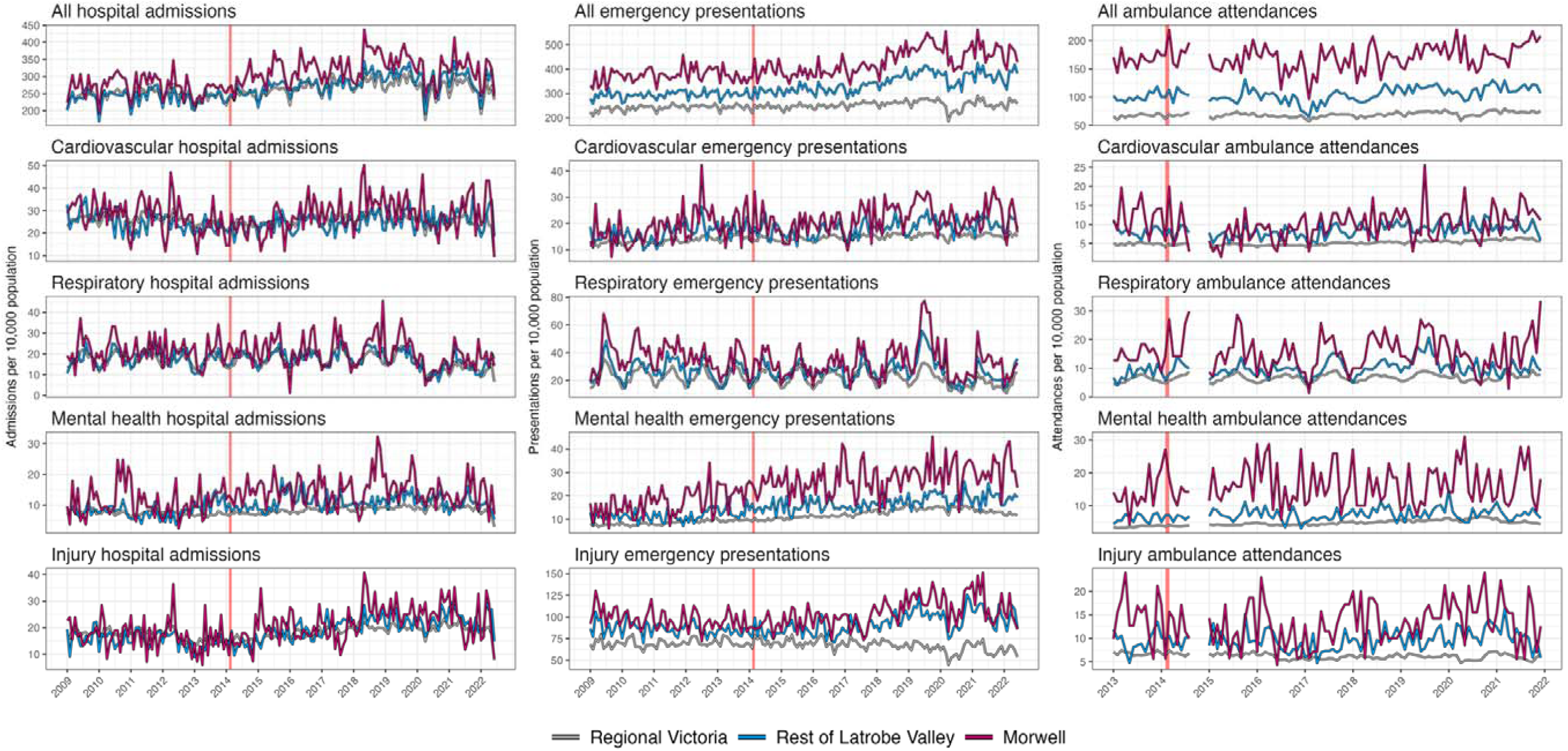
Crude time series plots indicating total monthly hospital admissions, emergency department presentations, and ambulance attendances, by condition, AMONG MEN.

### 3 Results tables

**Table S1.**
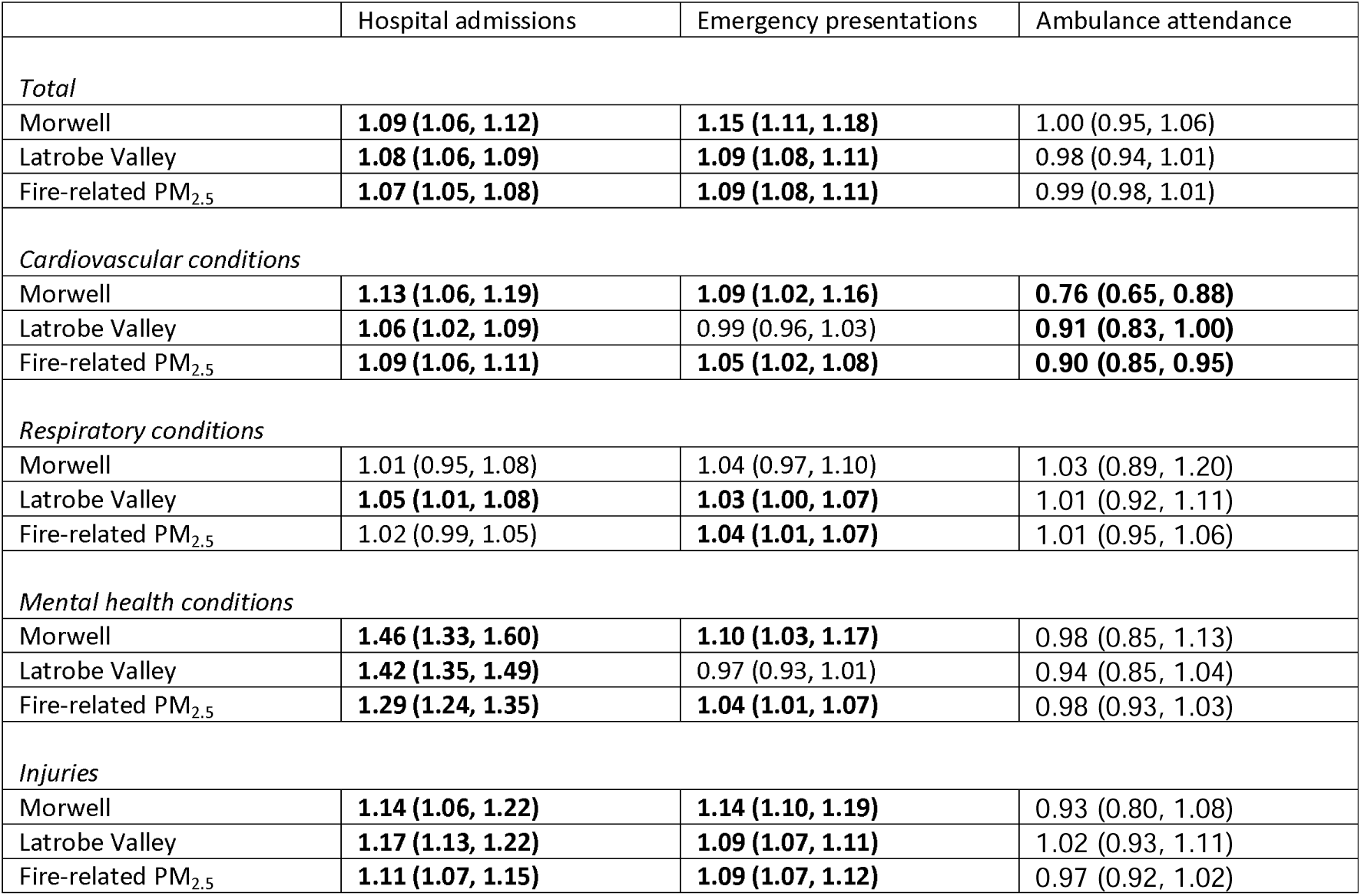
Changes in hospital admission, emergency presentations, and ambulance attendance in the eight years following the Hazelwood coalmine fire, relative risk (95% confidence interval) compared to the rest of regional Victoria.

**Table S2.**
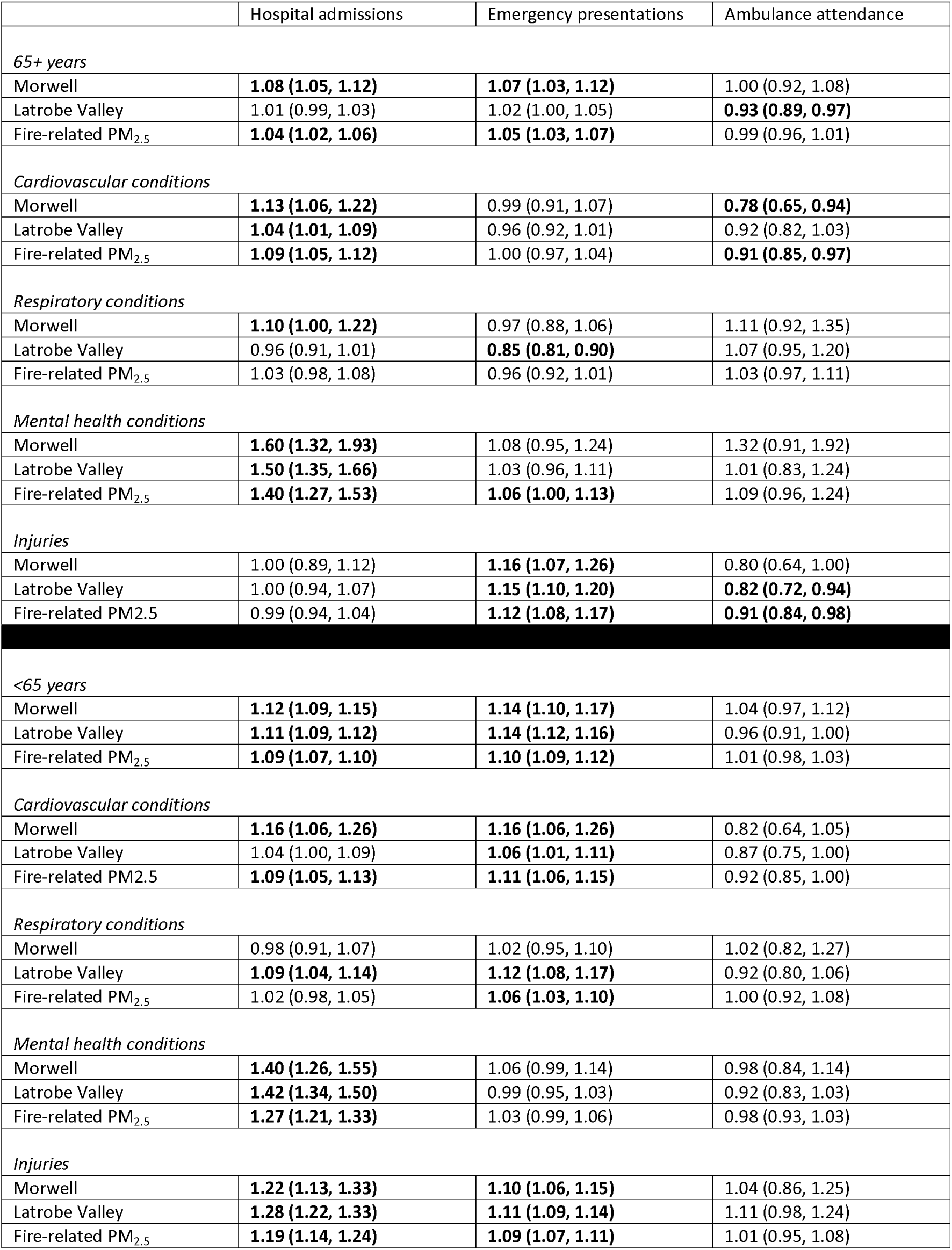
Changes in hospital admission, emergency presentations, and ambulance attendance in the eight years following the Hazelwood coalmine fire: by age group (65+, <65 years)

**Table S3.**
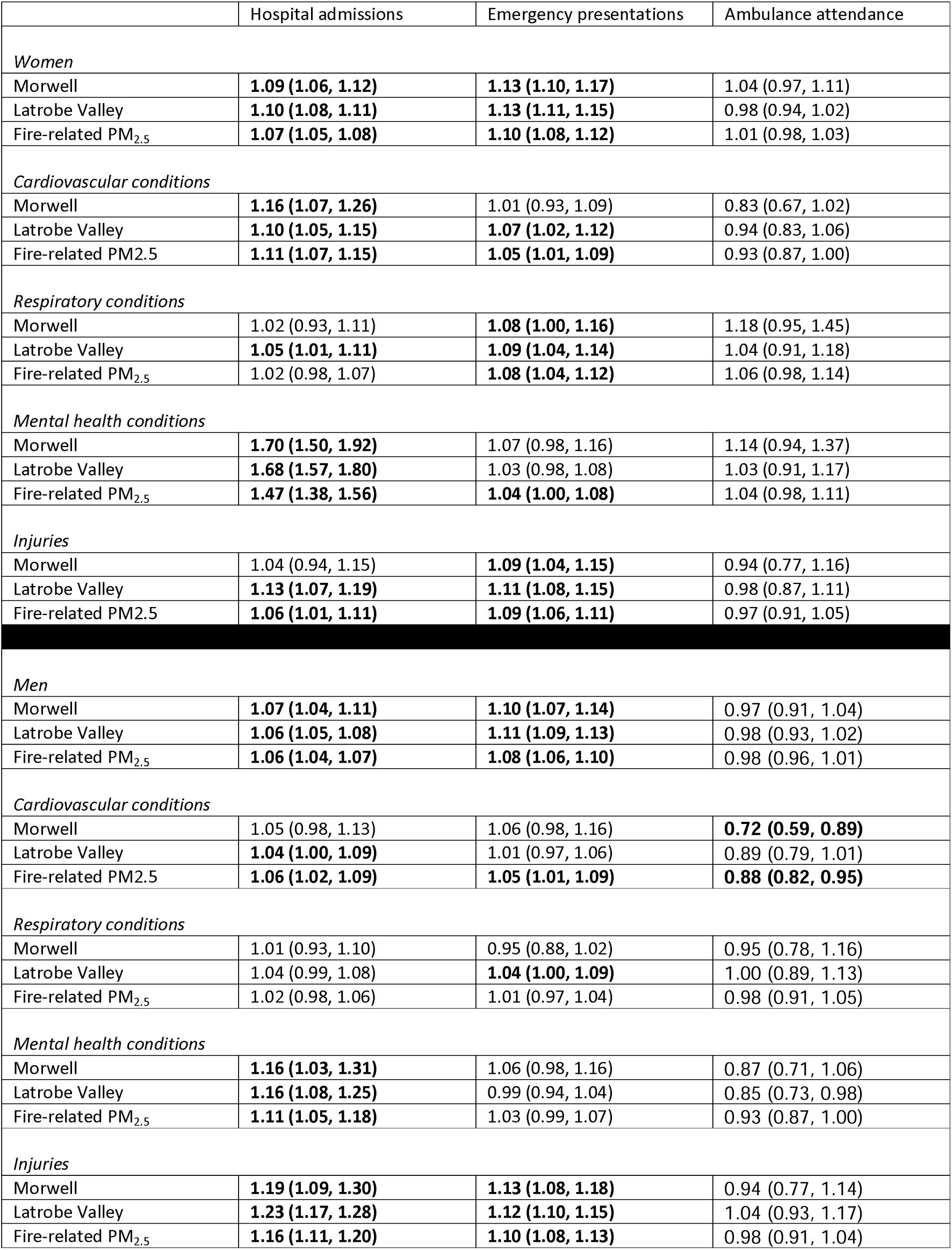
Changes in hospital admission, emergency presentations, and ambulance attendance in the eight years following the Hazelwood coalmine fire: by sex (women, men)

### 4 Sensitivity analyses

In sensitivity analyses, we added background PM_2.5_ data (23) to our models. The aim was not to evaluate the effect of background PM_2.5_ but adjust for any potential confounding due to it. As indicated by Figures S8 and S9, this had no appreciable influence on fire-related PM_2.5_ effects. Surprisingly, background PM_2.5_ was associated with reduced overall hospital, emergency, and ambulance use, with mixed effects on specific conditions. As a substantial body of evidence links background PM_2.5_ to poorer health (54), our findings require some examination.

**Figure S8.**
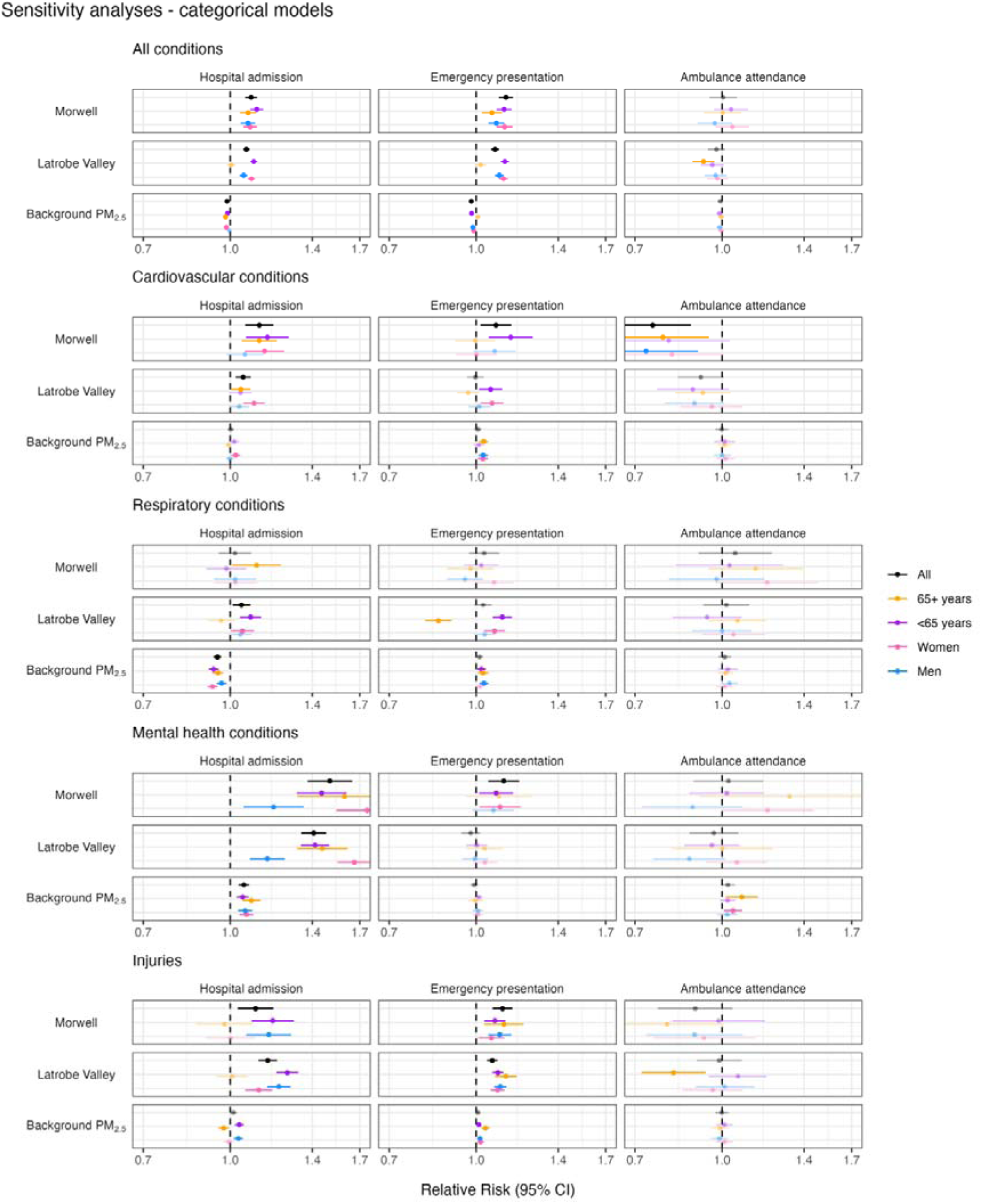
SENSITIVITY ANALYSIS ADDING BACKGROUND PM_2.5_: Changes in hospital admission, emergency presentations, and ambulance attendance in the eight years following the Hazelwood coalmine fire; categorical model (Morwell and rest of Latrobe Valley compared to rest of regional Victoria)

**Figure S9.**
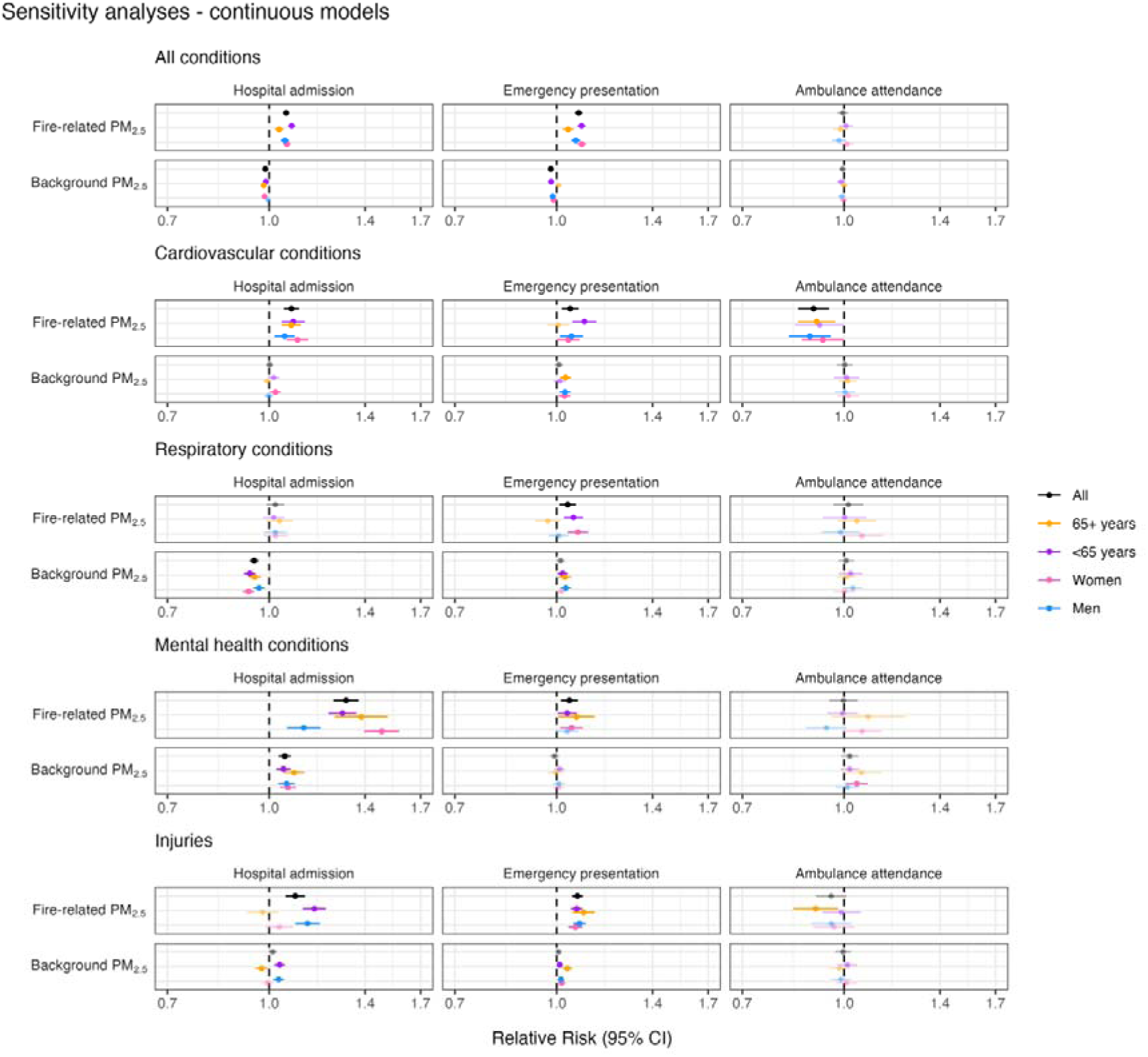
SENSITIVITY ANALYSIS ADDING BACKGROUND PM_2.5_: Changes in hospital admission, emergency presentations, and ambulance attendance in the eight years following the Hazelwood coalmine fire; continuous model (fire-related PM_2.5_ at Statistical Area level 2)

**Table S4.**
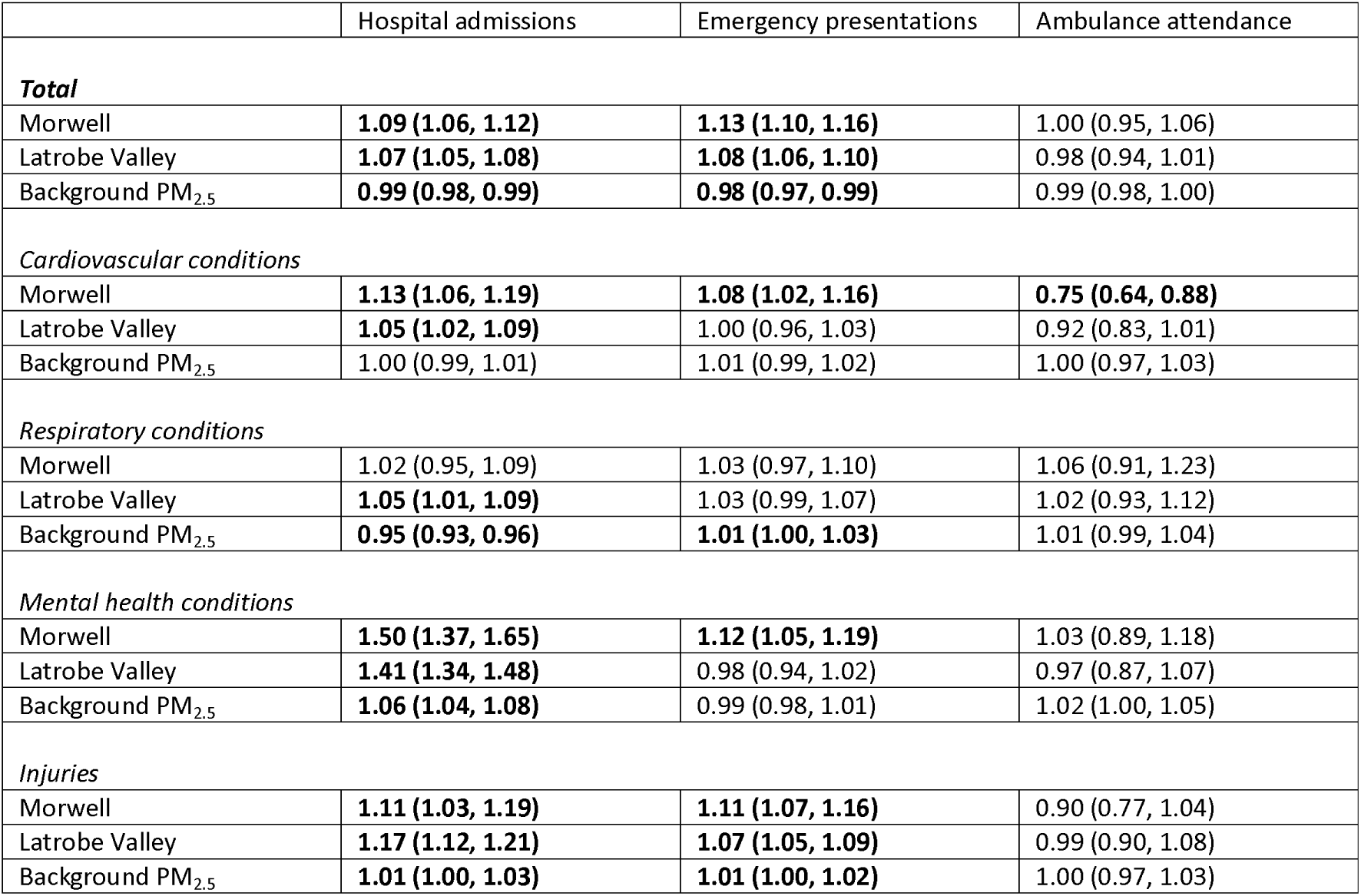
SENSITIVITY ANALYSIS ADDING BACKGROUND PM_2.5_: Changes in hospital admission, emergency presentations, and ambulance attendance in the eight years following the Hazelwood coalmine fire; categorical model (Morwell and rest of Latrobe Valley compared to rest of regional Victoria)

**Table S5.**
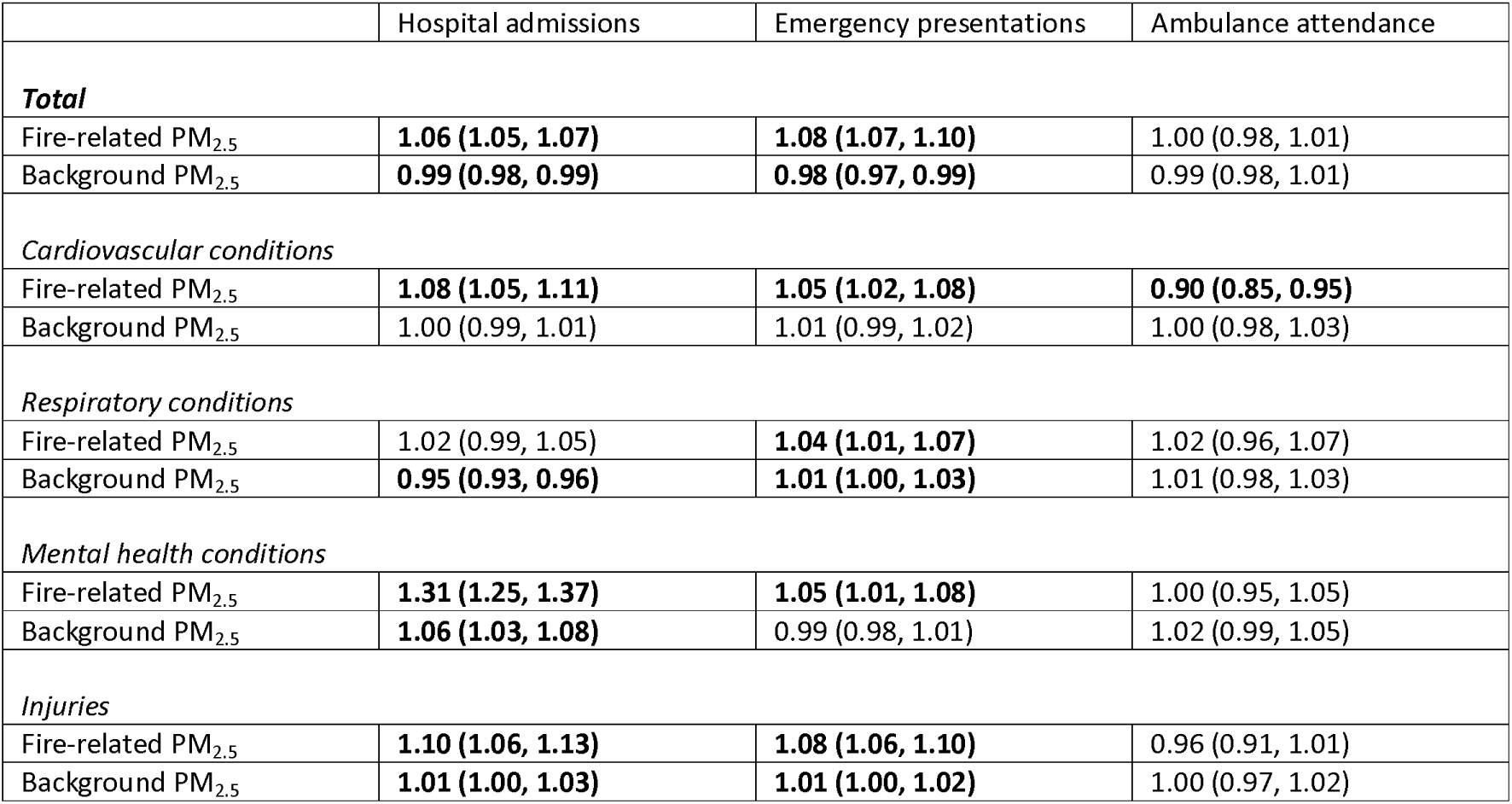
SENSITIVITY ANALYSIS ADDING BACKGROUND PM_2.5_: Changes in hospital admission, emergency presentations, and ambulance attendance in the eight years following the Hazelwood coalmine fire; continuous model (fire-related PM_2.5_ at Statistical Area level 2)

**Table S6.**
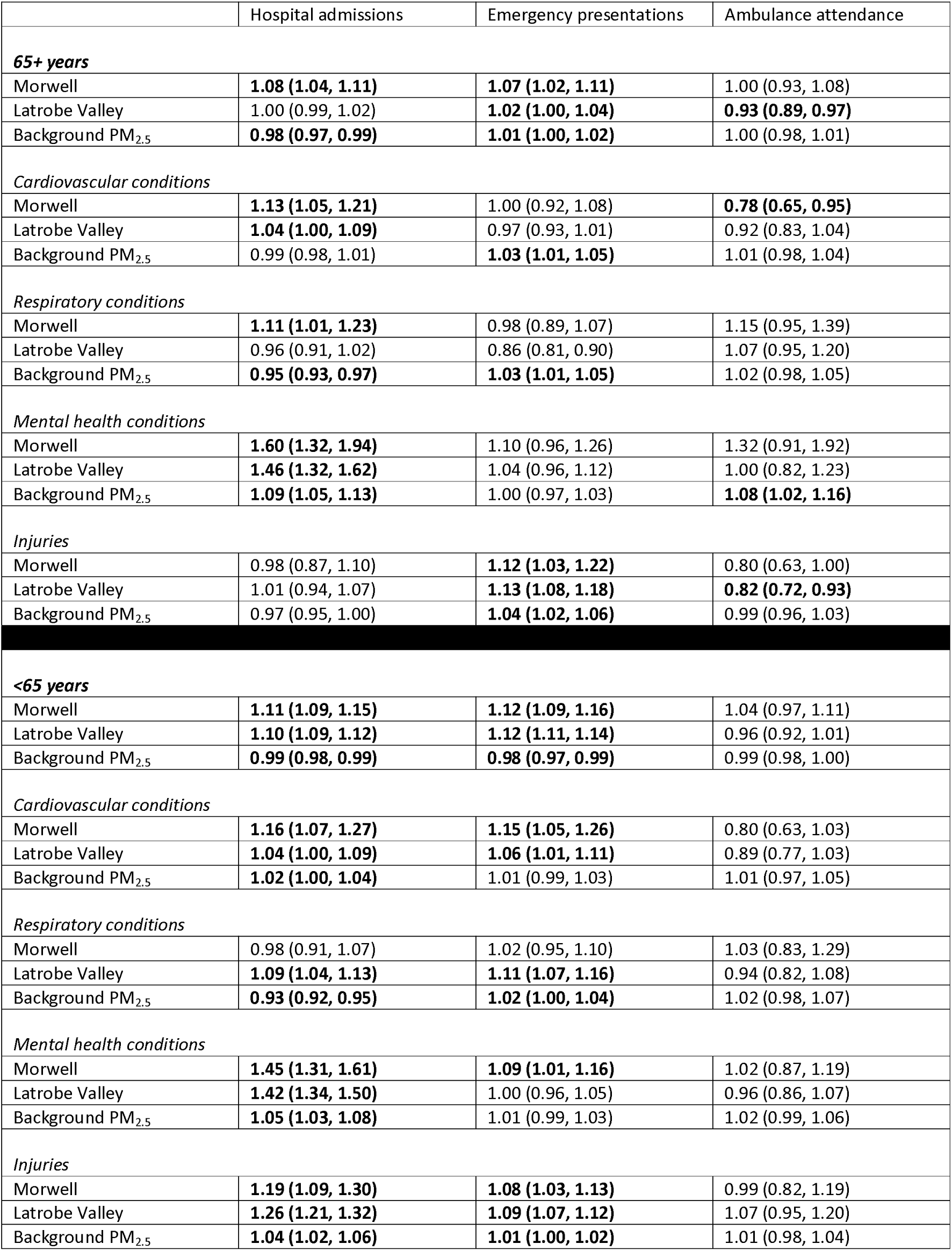
SENSITIVITY ANALYSIS ADDING BACKGROUND PM_2.5_: Changes in hospital admission, emergency presentations, and ambulance attendance in the eight years following the Hazelwood coalmine fire; categorical model (Morwell and rest of Latrobe Valley compared to rest of regional Victoria)

**Table S7.**
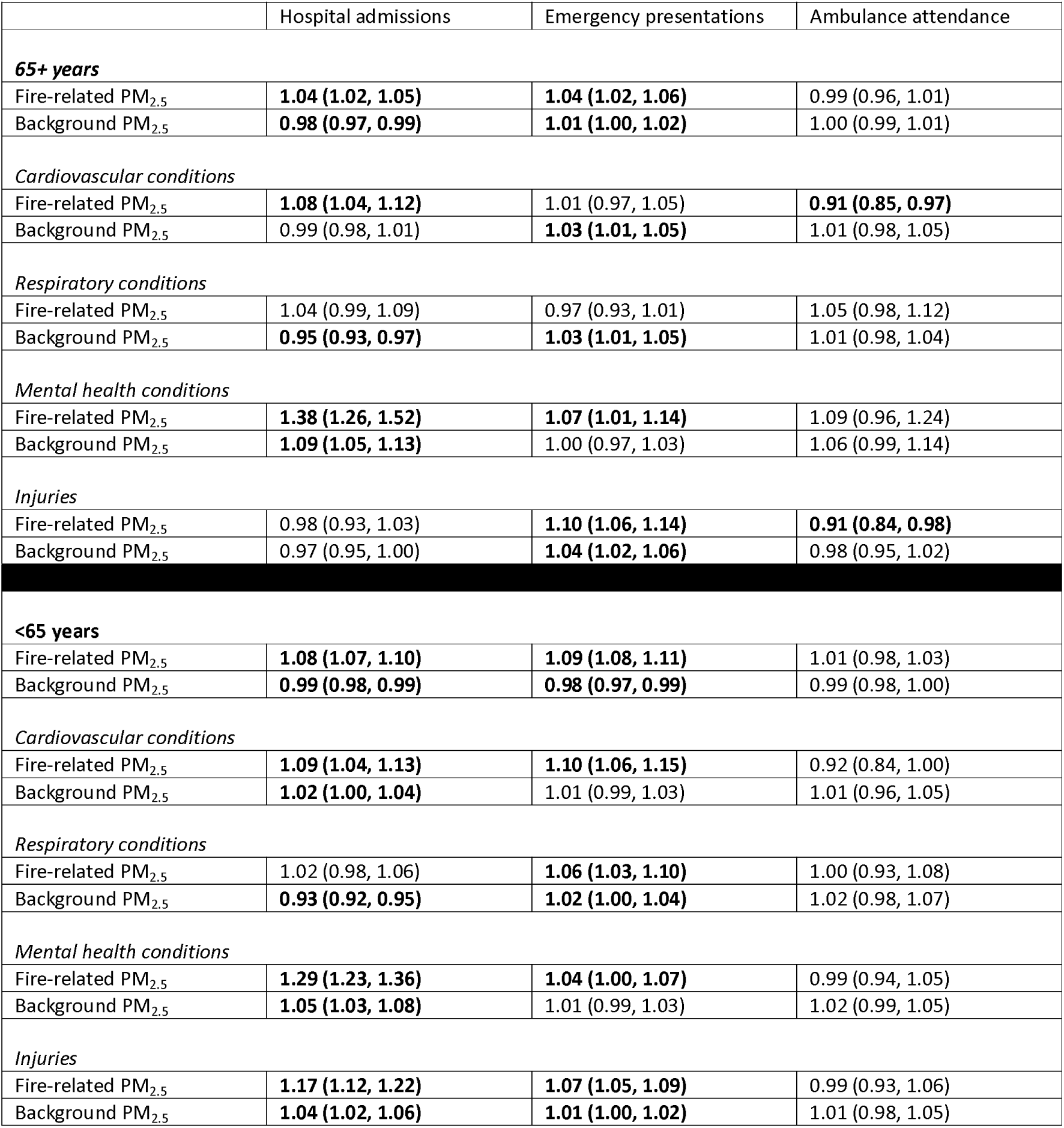
SENSITIVITY ANALYSIS ADDING BACKGROUND PM_2.5_: Changes in hospital admission, emergency presentations, and ambulance attendance in the eight years following the Hazelwood coalmine fire: by age group (65+, <65 years); continuous model (fire-related PM_2.5_ at Statistical Area level 2)

**Table S8.**
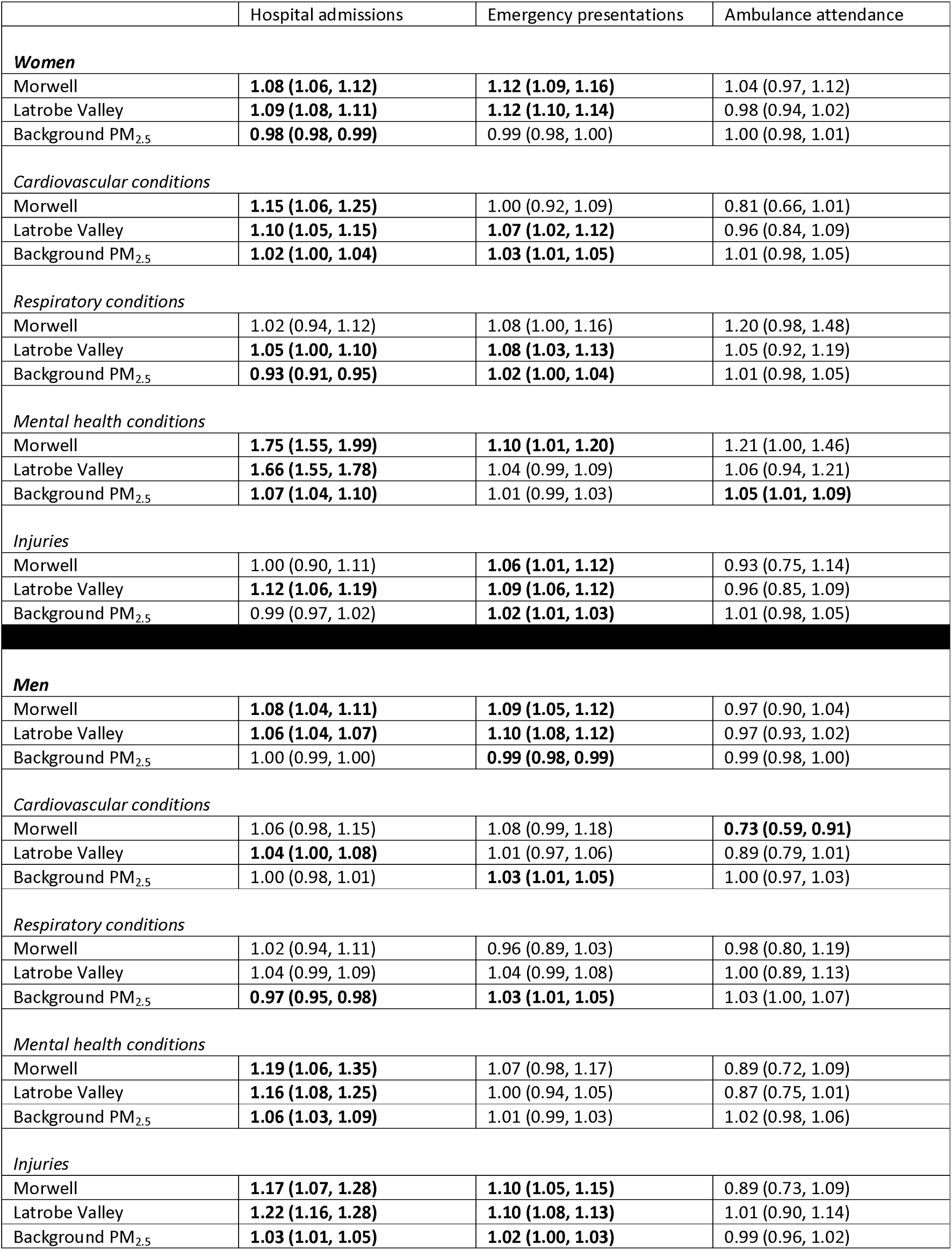
SENSITIVITY ANALYSIS ADDING BACKGROUND PM_2.5_: Changes in hospital admission, emergency presentations, and ambulance attendance in the eight years following the Hazelwood coalmine fire: by sex (women, men); categorical model (Morwell and rest of Latrobe Valley compared to rest of regional Victoria)

**Table S9.**
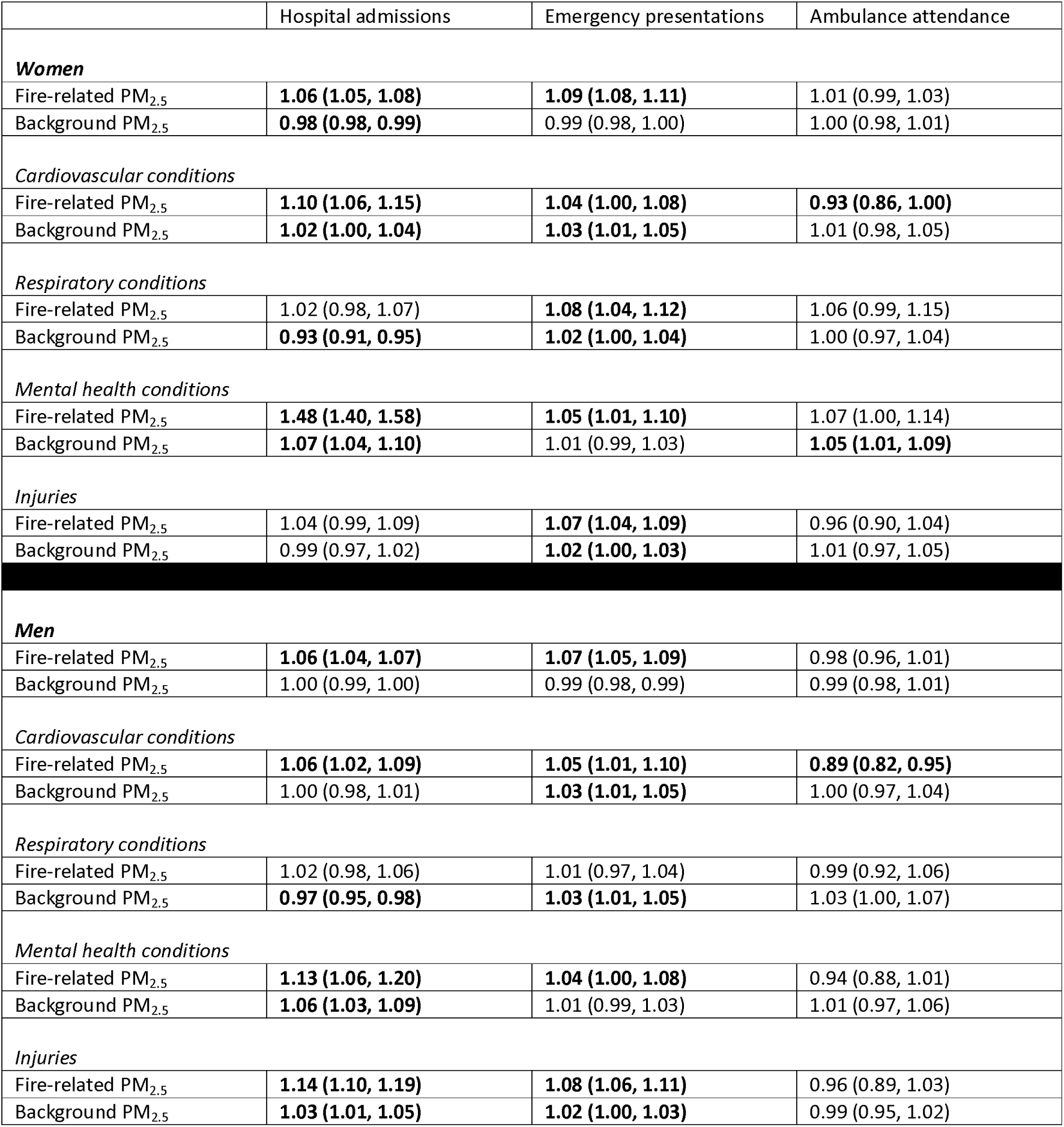
SENSITIVITY ANALYSIS ADDING BACKGROUND PM_2.5_: Changes in hospital admission, emergency presentations, and ambulance attendance in the eight years following the Hazelwood coalmine fire: by sex (women, men) ; continuous model (fire-related PM_2.5_ at Statistical Area level 2)

#### 4.1 Socioeconomic variance in the Latrobe Valley

The map in Figure S10 illustrates the distribution in socioeconomic deprivation between Morwell and the rest of the Latrobe Valley based on Index of Relative Socio-economic Advantage and Disadvantage (IRSAD) in 2021 (34). Morwell is in the 2^nd^ percentile for Victoria, indicating it is one of the most deprived areas in the state. The next lowest in the Latrobe Valley is Moe/Newborough, which is in the 5^th^ percentile. The remaining areas range between the 21^st^ and 42^nd^ percentiles (Churchill: 27^th^ , Traralgon [East]: 21^st^ , Traralgon [West]: 42^nd^ , Yallourn North/Glengarry: 30^th^ ).

**Figure S10.**
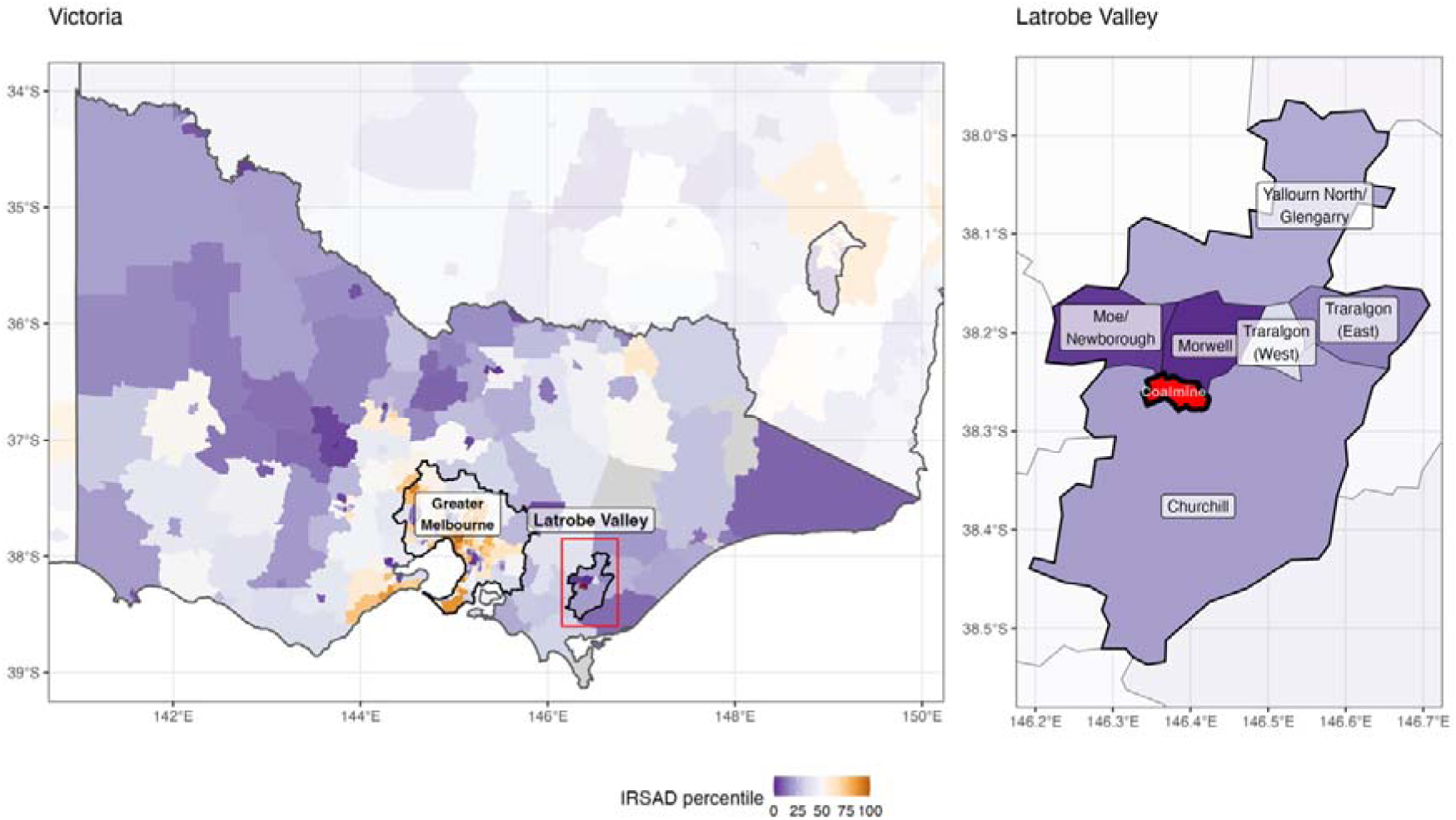
Map of Victorian socioeconomic (IRSAD) rankings in the Latrobe Valley.

#### 4.2 Examining why background PM_2.5_ was often associated with reduced service use

We suspected the effect may have been due to complex associations between air pollution and socioeconomic status. For instance, Figure S3 presents the correlation between Index of Relative Socioeconomic Advantage and Disadvantage (IRSAD) (34) and background PM_2.5_ (23) at Statistical Area Level 2. Both datasets were from 2021 to account for events that may have affected Morwell’s socioeconomic standing, namely the closure of the coalmine and power plant in 2017. These analyses are presented in Figure S3.

When taken as a whole, more advantaged areas in Victoria have higher levels of background PM_2.5_, likely due to greater amounts vehicle traffic and industrial activity; a standard deviation increase in the score (indicating greater advantage) was associated with an 8.3% (95%CI: 7.0% to 9.4%) increase in background PM_2.5_; this is reflected by the grey line in Figure S3. However, there were clear differences between Greater Melbourne (blue dots), in which background PM_2.5_ levels were generally higher and indicative of a positive association with socioeconomic status and regional Victoria (orange dots and line), where there was no detectable association (-0.9%, 95%CI: -2.1% to 0.3%).

**Figure S11.**
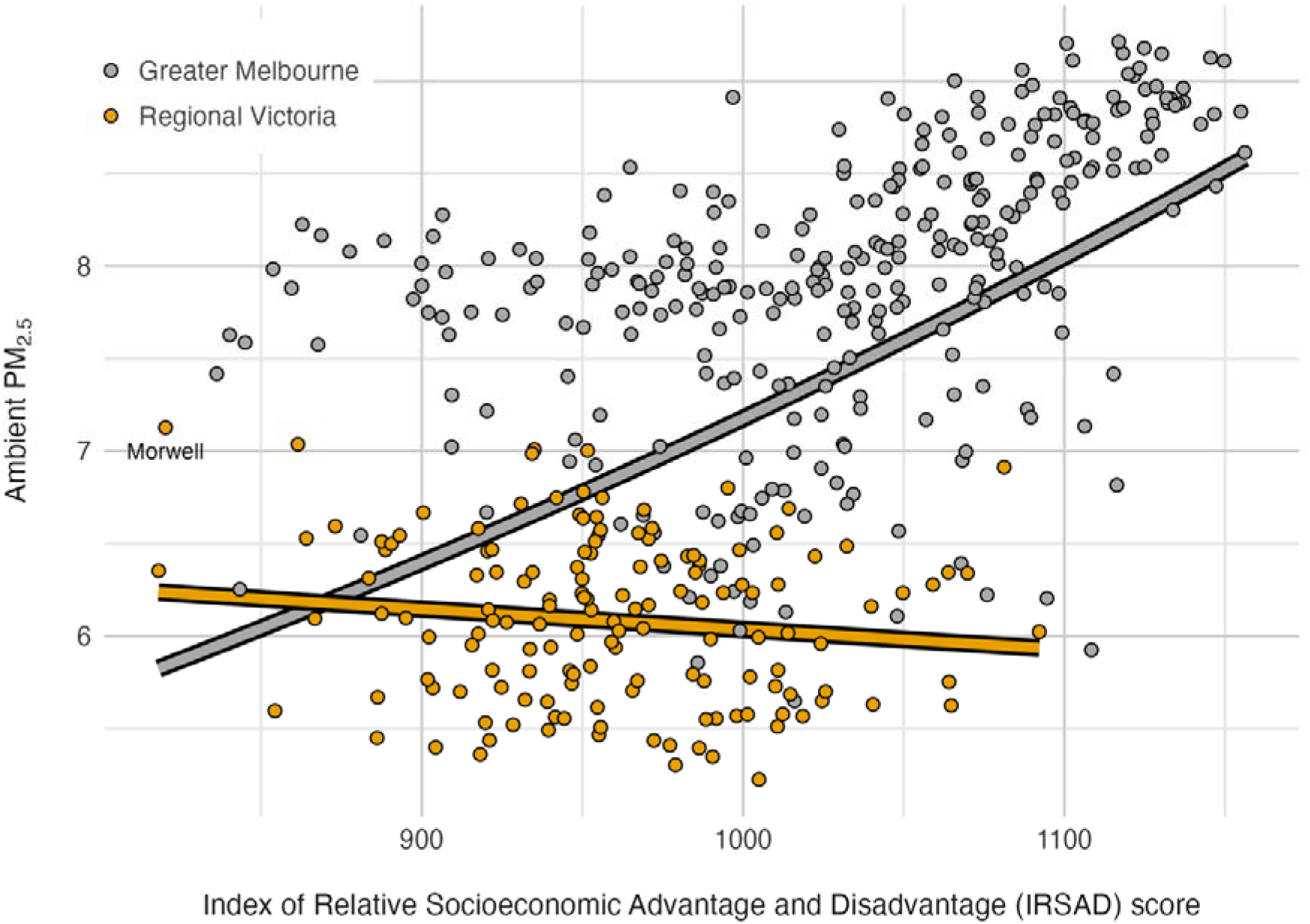
Relationship between socioeconomic status (based on IRSAD) and ambient PM_2.5_.

Another potential explanation for the unexpected negative associations between background PM_2.5_ and health service use is aggregation error. In other words, the overall level of background PM_2.5_ across Statistical Areas may poorly reflect actual exposures of people in a given area, since they ignore heterogeneity, particularly between populated areas with more vehicular/industrial activity and less populated areas. This problem is exaggerated in regional areas, since Statistical Areas are population-defined, using sizes that are designed to be roughly comparable, ranging from 3,000 to 25,000 people but averaging around 10,000 (17). This means they are geographically larger than urban Statistical Areas and background PM_2.5_ estimates at this level are less accurate. Further, as they have considerably more unpopulated space, their estimates are further biased downwards.

