## Supplementary materials for "Long-term effects of a coalmine fire on hospital and ambulance use: an interrupted time series study"

#### Table of Contents

*1 Fire-related PM$_{2.5}$ ... 3*

*2 Time series plots ... 5*

*3 Results tables ... 10*

*4 Sensitivity analyses ... 13*

4.1 Socioeconomic variance in the Latrobe Valley ... 21

4.2 Background PM$_{2.5}$ often associated with reduced service use ... 22

### 1 Fire-related $PM_{2.5}$

In the main document's *Introduction*, we compared daily mean fire-related $PM_{2.5}$ in Morwell (32.8 μg/m$^3$) and the rest of the Latrobe Valley (3.1 μg/m$^3$) using the "peak fire period", which we define as 9 February 2014 to 6 March 2014. Using modelled $PM_{2.5}$ data from Luhar et al. 2020 (1), this has been illustrated in Figure S1 (2). The area shaded pink highlights the timeframe in which there were clearly visible peaks in fire-related $PM_{2.5}$. Yet from 7 March 2014 onward, model suggests very low amounts of fire-related $PM_{2.5}$.

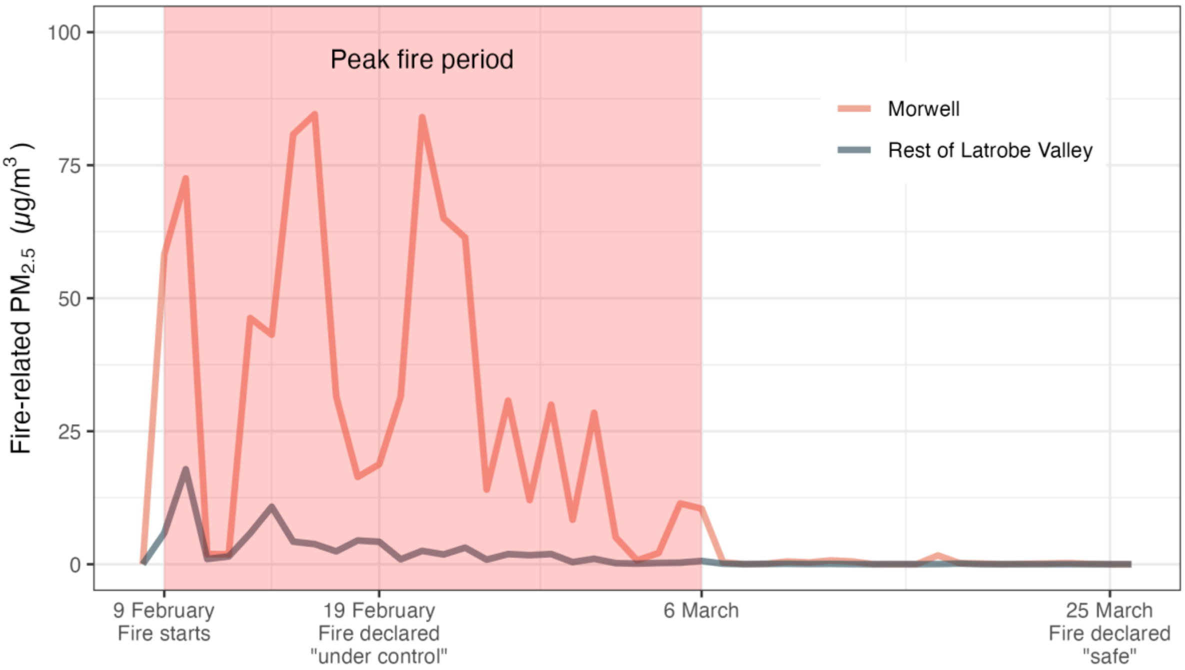

**Figure S1. Daily mean fire-related $PM_{2.5}$ during the Hazelwood coalmine fire in Morwell and the rest of the Latrobe Valley**

Figure S2 builds on the map in Figure 1 and the time series in Figure S1 to illustrate how much greater smoke exposure from the Hazelwood coalmine fire was in Morwell and – to a lesser extent – the Latrobe Valley. This figure uses a longer time frame for which fire-related $PM_{2.5}$ was estimated (2 February 2014 to 28 March 2014). Each line underneath the distribution curve stands in for a Statistical Area at Level 2. Morwell, which is red, is far to the right, indicating extreme amounts of cumulative daily mean fire-related $PM_{2.5}$. The rest of the Latrobe Valley, in yellow, follows, though distantly.

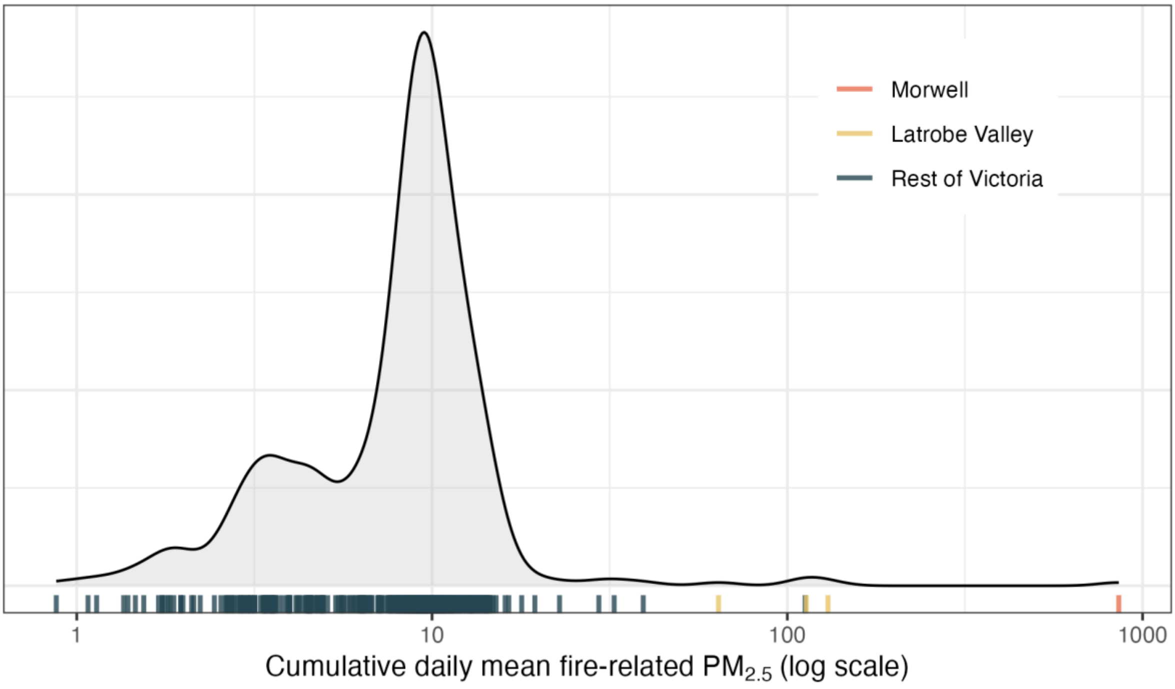

**Figure S2. Distributions of cumulative daily mean coalmine fire-related $PM_{2.5}$ by Statistical Area at Level 2**

#### 2 Time series plots

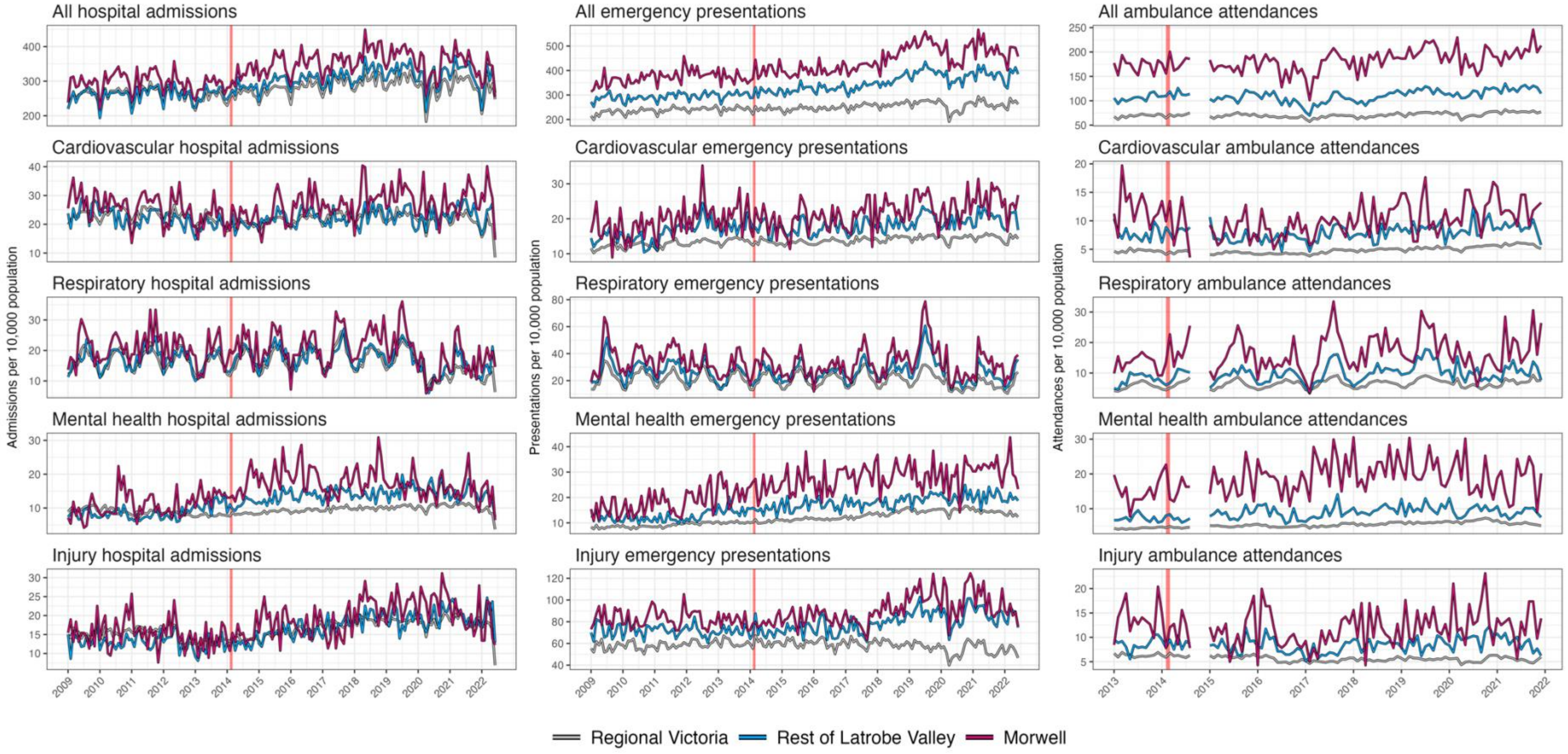

**Figure S3. Crude time series plots indicating total monthly hospital admissions, emergency department presentations, and ambulance attendances, by condition, TOTAL**

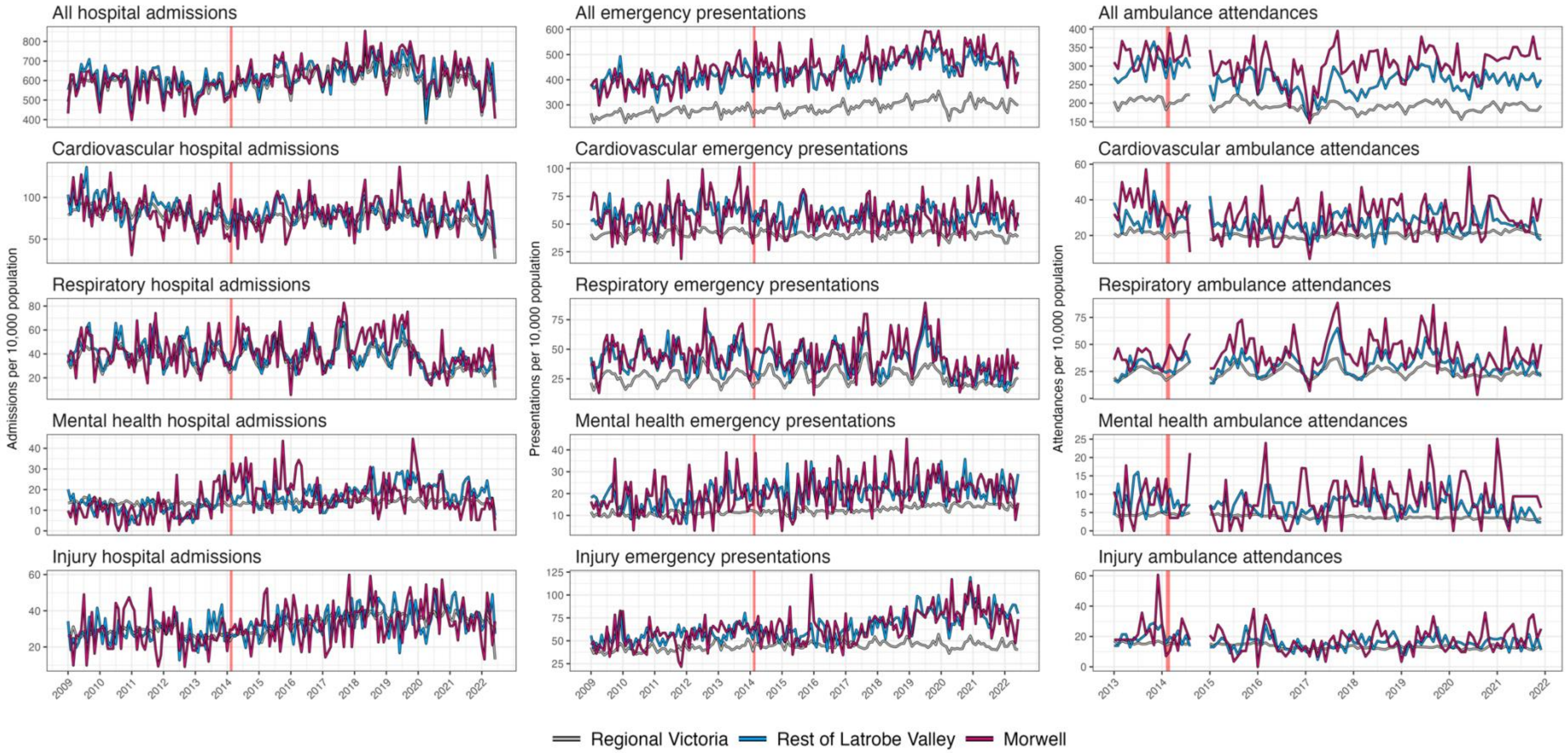

**Figure S4. Crude time series plots indicating total monthly hospital admissions, emergency department presentations, and ambulance attendances, by condition, AMONG THOSE AGED 65+ YEARS**

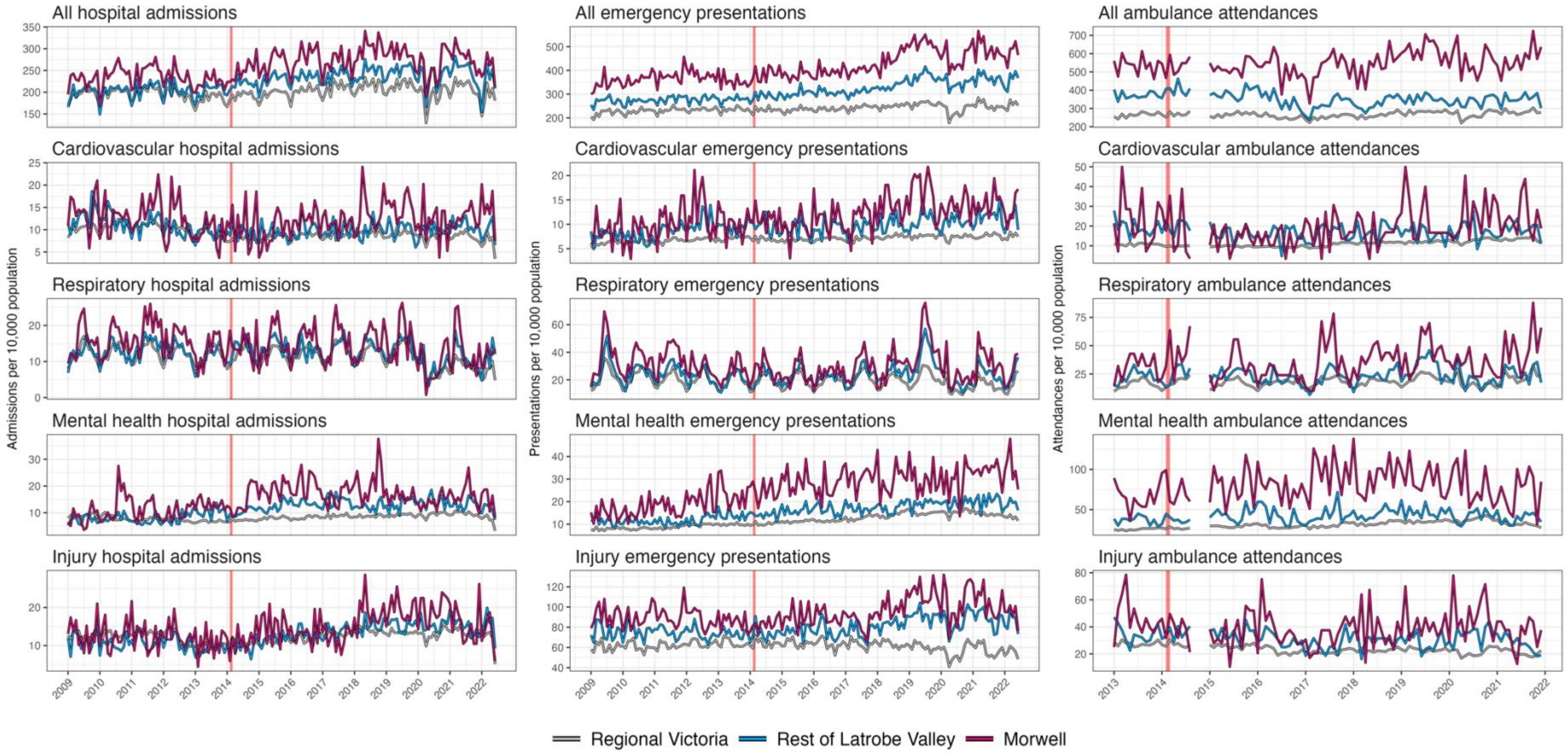

**Figure S5. Crude time series plots indicating total monthly hospital admissions, emergency department presentations, and ambulance attendances, by condition, AMONG THOSE AGED <65 YEARS**

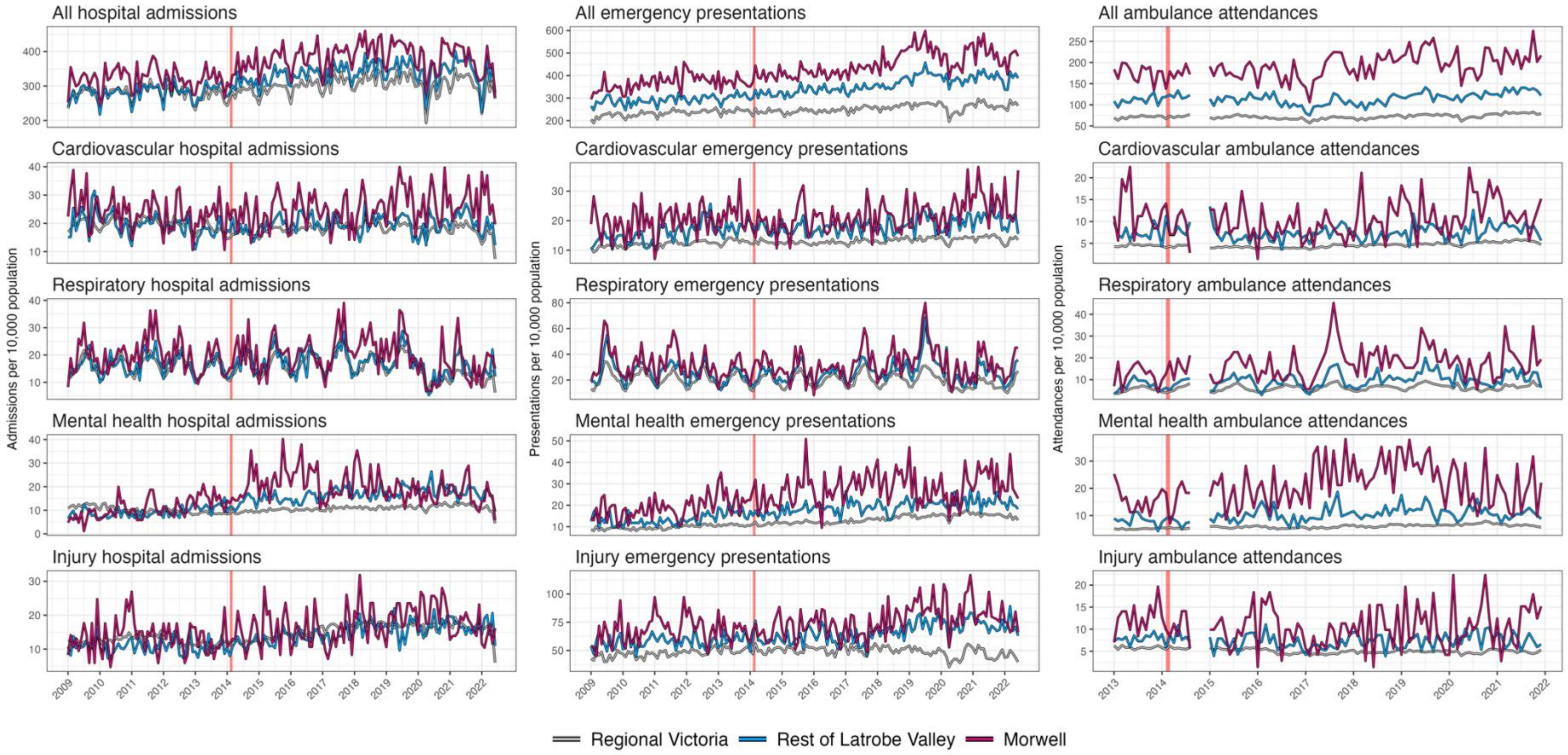

**Figure S6. Crude time series plots indicating total monthly hospital admissions, emergency department presentations, and ambulance attendances, by condition, AMONG WOMEN**

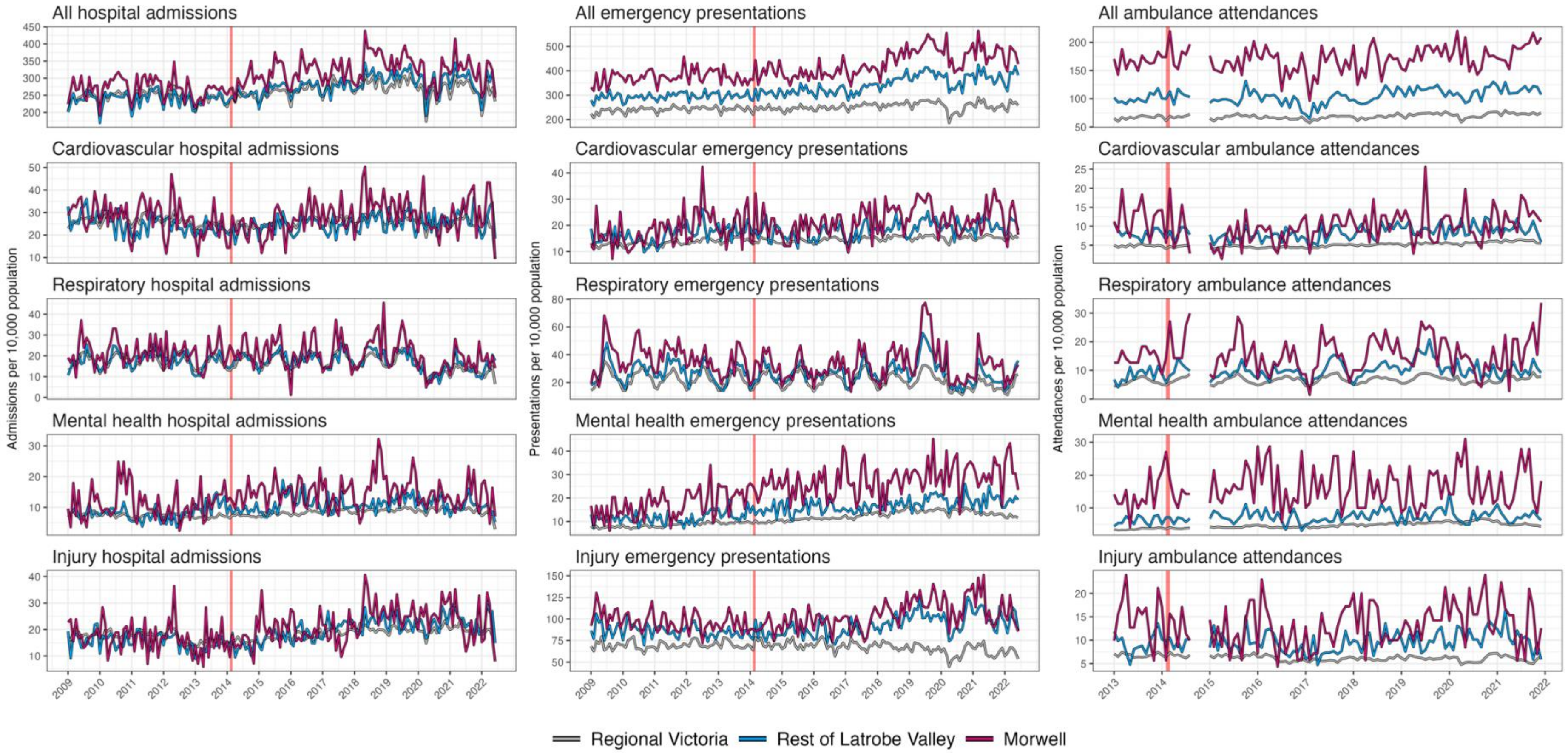

**Figure S7. Crude time series plots indicating total monthly hospital admissions, emergency department presentations, and ambulance attendances, by condition, AMONG MEN**

### 3 Results tables

**Table S1. Changes in hospital admission, emergency presentations, and ambulance attendance in the eight years following the Hazelwood coalmine fire, relative risk (95% confidence interval) compared to the rest of regional Victoria**

| | Hospital admissions | Emergency presentations | Ambulance attendance |
|---|---|---|---|
| *Total* | | | |
| Morwell | **1.09 (1.06, 1.12)** | **1.15 (1.11, 1.18)** | 1.00 (0.95, 1.06) |
| Latrobe Valley | **1.08 (1.06, 1.09)** | **1.09 (1.08, 1.11)** | 0.98 (0.94, 1.01) |
| Fire-related $PM_{2.5}$ | **1.07 (1.05, 1.08)** | **1.09 (1.08, 1.11)** | 0.99 (0.98, 1.01) |
| *Cardiovascular conditions* | | | |
| Morwell | **1.13 (1.06, 1.19)** | **1.09 (1.02, 1.16)** | **0.76 (0.65, 0.88)** |
| Latrobe Valley | **1.06 (1.02, 1.09)** | 0.99 (0.96, 1.03) | **0.91 (0.83, 1.00)** |
| Fire-related $PM_{2.5}$ | **1.09 (1.06, 1.11)** | **1.05 (1.02, 1.08)** | **0.90 (0.85, 0.95)** |
| *Respiratory conditions* | | | |
| Morwell | 1.01 (0.95, 1.08) | 1.04 (0.97, 1.10) | 1.03 (0.89, 1.20) |
| Latrobe Valley | **1.05 (1.01, 1.08)** | **1.03 (1.00, 1.07)** | 1.01 (0.92, 1.11) |
| Fire-related $PM_{2.5}$ | 1.02 (0.99, 1.05) | **1.04 (1.01, 1.07)** | 1.01 (0.95, 1.06) |
| *Mental health conditions* | | | |
| Morwell | **1.46 (1.33, 1.60)** | **1.10 (1.03, 1.17)** | 0.98 (0.85, 1.13) |
| Latrobe Valley | **1.42 (1.35, 1.49)** | 0.97 (0.93, 1.01) | 0.94 (0.85, 1.04) |
| Fire-related $PM_{2.5}$ | **1.29 (1.24, 1.35)** | **1.04 (1.01, 1.07)** | 0.98 (0.93, 1.03) |
| *Injuries* | | | |
| Morwell | **1.14 (1.06, 1.22)** | **1.14 (1.10, 1.19)** | 0.93 (0.80, 1.08) |
| Latrobe Valley | **1.17 (1.13, 1.22)** | **1.09 (1.07, 1.11)** | 1.02 (0.93, 1.11) |
| Fire-related $PM_{2.5}$ | **1.11 (1.07, 1.15)** | **1.09 (1.07, 1.12)** | 0.97 (0.92, 1.02) |

#### Table S2. Changes in hospital admission, emergency presentations, and ambulance attendance in the eight years following the Hazelwood coalmine fire: by age group (65+, <65 years)

| | Hospital admissions | Emergency presentations | Ambulance attendance |
|---|---|---|---|
| *65+ years* | | | |
| Morwell | **1.08 (1.05, 1.12)** | **1.07 (1.03, 1.12)** | 1.00 (0.92, 1.08) |
| Latrobe Valley | 1.01 (0.99, 1.03) | 1.02 (1.00, 1.05) | **0.93 (0.89, 0.97)** |
| Fire-related $PM_{2.5}$ | **1.04 (1.02, 1.06)** | **1.05 (1.03, 1.07)** | 0.99 (0.96, 1.01) |
| *Cardiovascular conditions* | | | |
| Morwell | **1.13 (1.06, 1.22)** | 0.99 (0.91, 1.07) | **0.78 (0.65, 0.94)** |
| Latrobe Valley | **1.04 (1.01, 1.09)** | 0.96 (0.92, 1.01) | 0.92 (0.82, 1.03) |
| Fire-related $PM_{2.5}$ | **1.09 (1.05, 1.12)** | 1.00 (0.97, 1.04) | **0.91 (0.85, 0.97)** |
| *Respiratory conditions* | | | |
| Morwell | **1.10 (1.00, 1.22)** | 0.97 (0.88, 1.06) | 1.11 (0.92, 1.35) |
| Latrobe Valley | 0.96 (0.91, 1.01) | **0.85 (0.81, 0.90)** | 1.07 (0.95, 1.20) |
| Fire-related $PM_{2.5}$ | 1.03 (0.98, 1.08) | 0.96 (0.92, 1.01) | 1.03 (0.97, 1.11) |
| *Mental health conditions* | | | |
| Morwell | **1.60 (1.32, 1.93)** | 1.08 (0.95, 1.24) | 1.32 (0.91, 1.92) |
| Latrobe Valley | **1.50 (1.35, 1.66)** | 1.03 (0.96, 1.11) | 1.01 (0.83, 1.24) |
| Fire-related $PM_{2.5}$ | **1.40 (1.27, 1.53)** | **1.06 (1.00, 1.13)** | 1.09 (0.96, 1.24) |
| *Injuries* | | | |
| Morwell | 1.00 (0.89, 1.12) | **1.16 (1.07, 1.26)** | 0.80 (0.64, 1.00) |
| Latrobe Valley | 1.00 (0.94, 1.07) | **1.15 (1.10, 1.20)** | **0.82 (0.72, 0.94)** |
| Fire-related PM2.5 | 0.99 (0.94, 1.04) | **1.12 (1.08, 1.17)** | **0.91 (0.84, 0.98)** |
| *<65 years* | | | |
| Morwell | **1.12 (1.09, 1.15)** | **1.14 (1.10, 1.17)** | 1.04 (0.97, 1.12) |
| Latrobe Valley | **1.11 (1.09, 1.12)** | **1.14 (1.12, 1.16)** | 0.96 (0.91, 1.00) |
| Fire-related $PM_{2.5}$ | **1.09 (1.07, 1.10)** | **1.10 (1.09, 1.12)** | 1.01 (0.98, 1.03) |
| *Cardiovascular conditions* | | | |
| Morwell | **1.16 (1.06, 1.26)** | **1.16 (1.06, 1.26)** | 0.82 (0.64, 1.05) |
| Latrobe Valley | 1.04 (1.00, 1.09) | **1.06 (1.01, 1.11)** | 0.87 (0.75, 1.00) |
| Fire-related PM2.5 | **1.09 (1.05, 1.13)** | **1.11 (1.06, 1.15)** | 0.92 (0.85, 1.00) |
| *Respiratory conditions* | | | |
| Morwell | 0.98 (0.91, 1.07) | 1.02 (0.95, 1.10) | 1.02 (0.82, 1.27) |
| Latrobe Valley | **1.09 (1.04, 1.14)** | **1.12 (1.08, 1.17)** | 0.92 (0.80, 1.06) |
| Fire-related $PM_{2.5}$ | 1.02 (0.98, 1.05) | **1.06 (1.03, 1.10)** | 1.00 (0.92, 1.08) |
| *Mental health conditions* | | | |
| Morwell | **1.40 (1.26, 1.55)** | 1.06 (0.99, 1.14) | 0.98 (0.84, 1.14) |
| Latrobe Valley | **1.42 (1.34, 1.50)** | 0.99 (0.95, 1.03) | 0.92 (0.83, 1.03) |
| Fire-related $PM_{2.5}$ | **1.27 (1.21, 1.33)** | 1.03 (0.99, 1.06) | 0.98 (0.93, 1.03) |
| *Injuries* | | | |
| Morwell | **1.22 (1.13, 1.33)** | **1.10 (1.06, 1.15)** | 1.04 (0.86, 1.25) |
| Latrobe Valley | **1.28 (1.22, 1.33)** | **1.11 (1.09, 1.14)** | 1.11 (0.98, 1.24) |
| Fire-related $PM_{2.5}$ | **1.19 (1.14, 1.24)** | **1.09 (1.07, 1.11)** | 1.01 (0.95, 1.08) |

#### Table S3. Changes in hospital admission, emergency presentations, and ambulance attendance in the eight years following the Hazelwood coalmine fire: by sex (women, men)

| | Hospital admissions | Emergency presentations | Ambulance attendance |
|---|---|---|---|
| *Women* | | | |
| Morwell | **1.09 (1.06, 1.12)** | **1.13 (1.10, 1.17)** | 1.04 (0.97, 1.11) |
| Latrobe Valley | **1.10 (1.08, 1.11)** | **1.13 (1.11, 1.15)** | 0.98 (0.94, 1.02) |
| Fire-related $PM_{2.5}$ | **1.07 (1.05, 1.08)** | **1.10 (1.08, 1.12)** | 1.01 (0.98, 1.03) |
| *Cardiovascular conditions* | | | |
| Morwell | **1.16 (1.07, 1.26)** | 1.01 (0.93, 1.09) | 0.83 (0.67, 1.02) |
| Latrobe Valley | **1.10 (1.05, 1.15)** | **1.07 (1.02, 1.12)** | 0.94 (0.83, 1.06) |
| Fire-related PM2.5 | **1.11 (1.07, 1.15)** | **1.05 (1.01, 1.09)** | 0.93 (0.87, 1.00) |
| *Respiratory conditions* | | | |
| Morwell | 1.02 (0.93, 1.11) | **1.08 (1.00, 1.16)** | 1.18 (0.95, 1.45) |
| Latrobe Valley | **1.05 (1.01, 1.11)** | **1.09 (1.04, 1.14)** | 1.04 (0.91, 1.18) |
| Fire-related $PM_{2.5}$ | 1.02 (0.98, 1.07) | **1.08 (1.04, 1.12)** | 1.06 (0.98, 1.14) |
| *Mental health conditions* | | | |
| Morwell | **1.70 (1.50, 1.92)** | 1.07 (0.98, 1.16) | 1.14 (0.94, 1.37) |
| Latrobe Valley | **1.68 (1.57, 1.80)** | 1.03 (0.98, 1.08) | 1.03 (0.91, 1.17) |
| Fire-related $PM_{2.5}$ | **1.47 (1.38, 1.56)** | **1.04 (1.00, 1.08)** | 1.04 (0.98, 1.11) |
| *Injuries* | | | |
| Morwell | 1.04 (0.94, 1.15) | **1.09 (1.04, 1.15)** | 0.94 (0.77, 1.16) |
| Latrobe Valley | **1.13 (1.07, 1.19)** | **1.11 (1.08, 1.15)** | 0.98 (0.87, 1.11) |
| Fire-related PM2.5 | **1.06 (1.01, 1.11)** | **1.09 (1.06, 1.11)** | 0.97 (0.91, 1.05) |
| *Men* | | | |
| Morwell | **1.07 (1.04, 1.11)** | **1.10 (1.07, 1.14)** | 0.97 (0.91, 1.04) |
| Latrobe Valley | **1.06 (1.05, 1.08)** | **1.11 (1.09, 1.13)** | 0.98 (0.93, 1.02) |
| Fire-related $PM_{2.5}$ | **1.06 (1.04, 1.07)** | **1.08 (1.06, 1.10)** | 0.98 (0.96, 1.01) |
| *Cardiovascular conditions* | | | |
| Morwell | 1.05 (0.98, 1.13) | 1.06 (0.98, 1.16) | **0.72 (0.59, 0.89)** |
| Latrobe Valley | **1.04 (1.00, 1.09)** | 1.01 (0.97, 1.06) | 0.89 (0.79, 1.01) |
| Fire-related PM2.5 | **1.06 (1.02, 1.09)** | **1.05 (1.01, 1.09)** | **0.88 (0.82, 0.95)** |
| *Respiratory conditions* | | | |
| Morwell | 1.01 (0.93, 1.10) | 0.95 (0.88, 1.02) | 0.95 (0.78, 1.16) |
| Latrobe Valley | 1.04 (0.99, 1.08) | **1.04 (1.00, 1.09)** | 1.00 (0.89, 1.13) |
| Fire-related $PM_{2.5}$ | 1.02 (0.98, 1.06) | 1.01 (0.97, 1.04) | 0.98 (0.91, 1.05) |
| *Mental health conditions* | | | |
| Morwell | **1.16 (1.03, 1.31)** | 1.06 (0.98, 1.16) | 0.87 (0.71, 1.06) |
| Latrobe Valley | **1.16 (1.08, 1.25)** | 0.99 (0.94, 1.04) | 0.85 (0.73, 0.98) |
| Fire-related $PM_{2.5}$ | **1.11 (1.05, 1.18)** | 1.03 (0.99, 1.07) | 0.93 (0.87, 1.00) |
| *Injuries* | | | |
| Morwell | **1.19 (1.09, 1.30)** | **1.13 (1.08, 1.18)** | 0.94 (0.77, 1.14) |
| Latrobe Valley | **1.23 (1.17, 1.28)** | **1.12 (1.10, 1.15)** | 1.04 (0.93, 1.17) |
| Fire-related $PM_{2.5}$ | **1.16 (1.11, 1.20)** | **1.10 (1.08, 1.13)** | 0.98 (0.91, 1.04) |

### 4 Sensitivity analyses

In sensitivity analyses, we added background $PM_{2.5}$ data (3) to our models. The aim was not to evaluate the effect of background $PM_{2.5}$ but adjust for any potential confounding due to it. As indicated by Figures S8 and S9, this had no appreciable influence on fire-related $PM_{2.5}$ effects. Surprisingly, background $PM_{2.5}$ was associated with reduced overall hospital, emergency, and ambulance use, with mixed effects on specific conditions. As a substantial body of evidence links background $PM_{2.5}$ to poorer health (4), our findings require some examination.

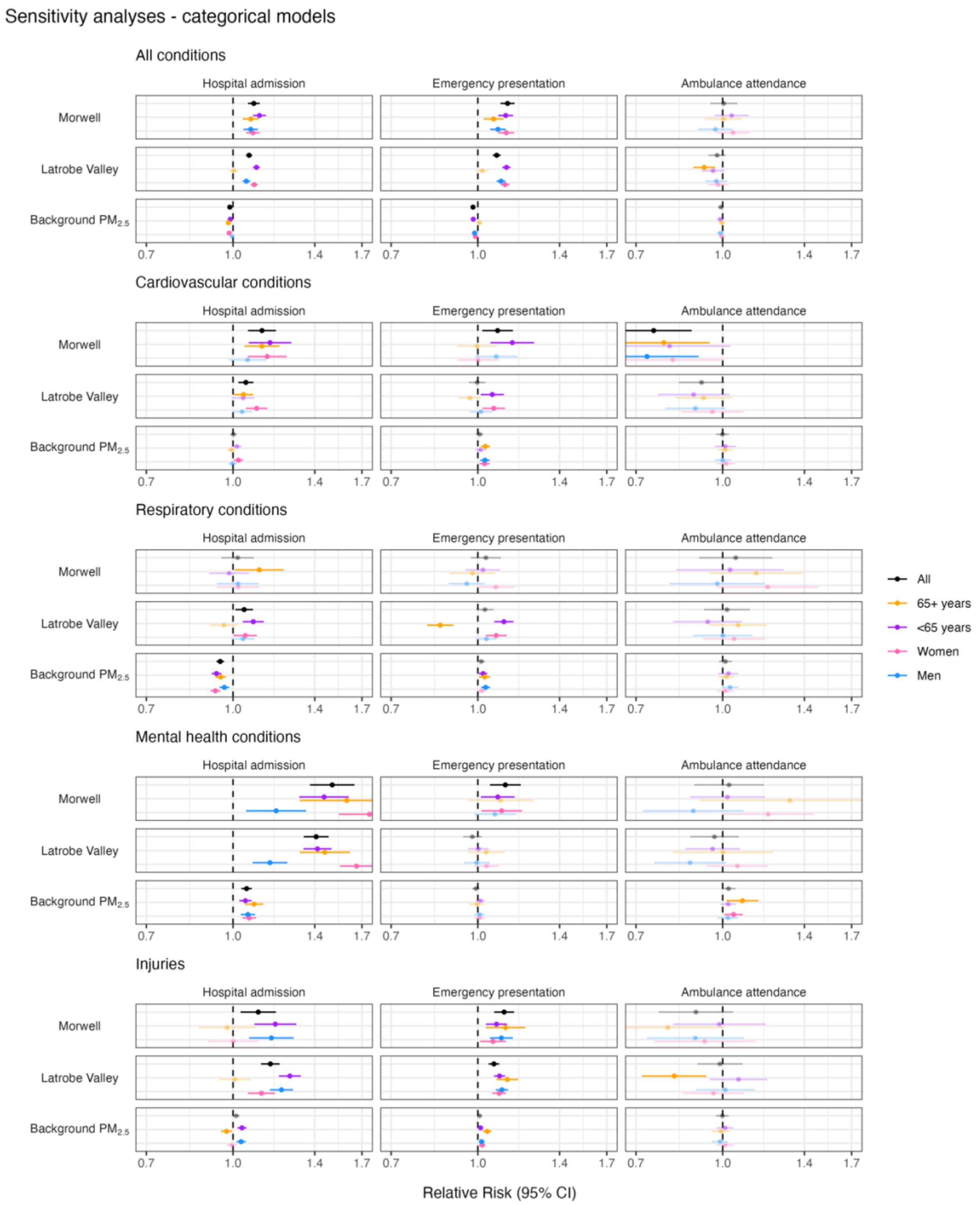

**Figure S8. SENSITIVITY ANALYSIS ADDING BACKGROUND $PM_{2.5}$: Changes in hospital admission, emergency presentations, and ambulance attendance in the eight years following the Hazelwood coalmine fire; categorical model (Morwell and rest of Latrobe Valley compared to rest of regional Victoria)**

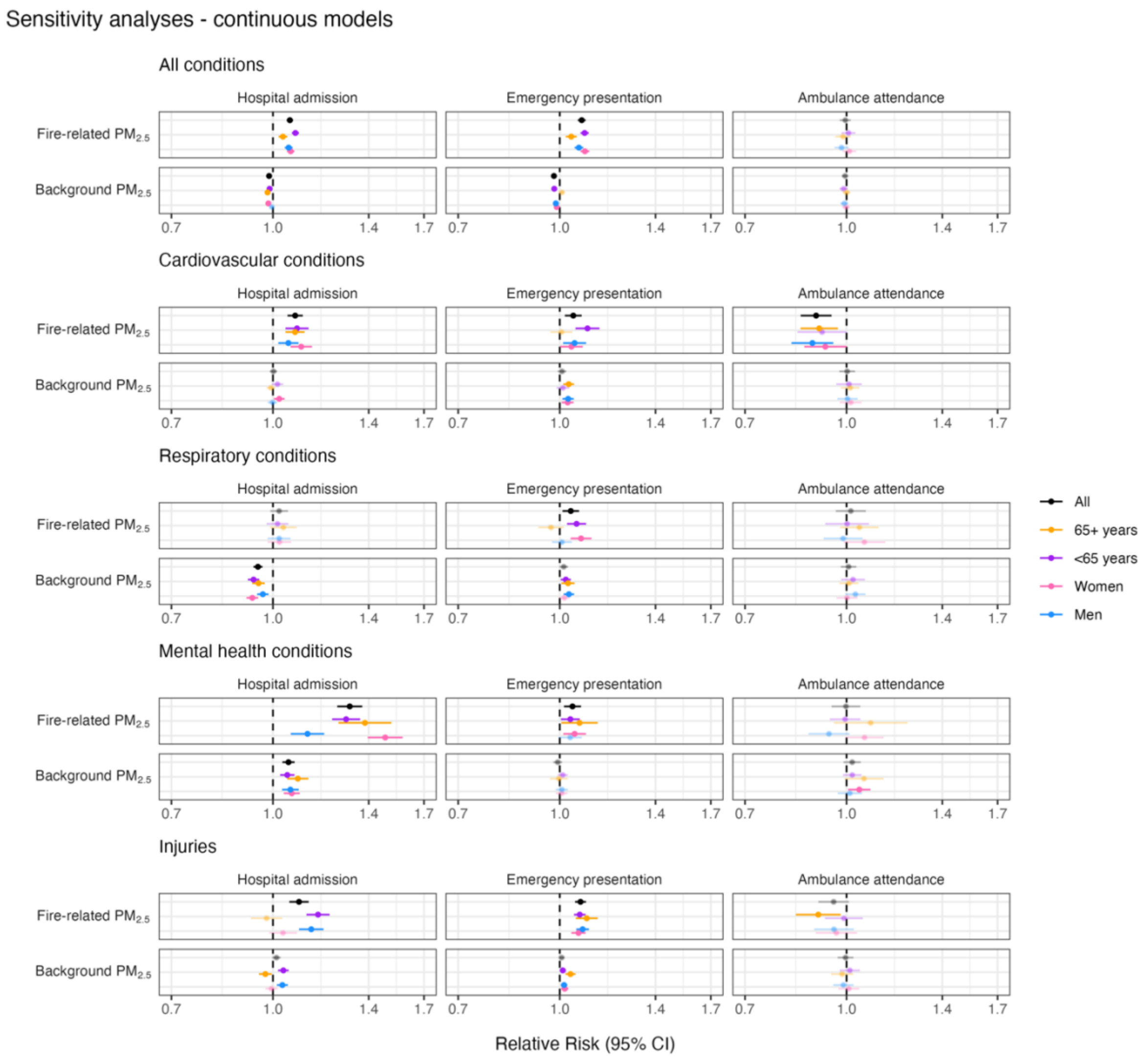

**Figure S9. SENSITIVITY ANALYSIS ADDING BACKGROUND $PM_{2.5}$: Changes in hospital admission, emergency presentations, and ambulance attendance in the eight years following the Hazelwood coalmine fire; continuous model (fire-related $PM_{2.5}$ at Statistical Area level 2)**

##### Table S4. SENSITIVITY ANALYSIS ADDING BACKGROUND PM$_{2.5}$: Changes in hospital admission, emergency presentations, and ambulance attendance in the eight years following the Hazelwood coalmine fire; categorical model (Morwell and rest of Latrobe Valley compared to rest of regional Victoria)

| | Hospital admissions | Emergency presentations | Ambulance attendance |
|---|---|---|---|
| ***Total*** | | | |
| Morwell | **1.09 (1.06, 1.12)** | **1.13 (1.10, 1.16)** | 1.00 (0.95, 1.06) |
| Latrobe Valley | **1.07 (1.05, 1.08)** | **1.08 (1.06, 1.10)** | 0.98 (0.94, 1.01) |
| Background PM$_{2.5}$ | **0.99 (0.98, 0.99)** | **0.98 (0.97, 0.99)** | 0.99 (0.98, 1.00) |
| *Cardiovascular conditions* | | | |
| Morwell | **1.13 (1.06, 1.19)** | **1.08 (1.02, 1.16)** | **0.75 (0.64, 0.88)** |
| Latrobe Valley | **1.05 (1.02, 1.09)** | 1.00 (0.96, 1.03) | 0.92 (0.83, 1.01) |
| Background PM$_{2.5}$ | 1.00 (0.99, 1.01) | 1.01 (0.99, 1.02) | 1.00 (0.97, 1.03) |
| *Respiratory conditions* | | | |
| Morwell | 1.02 (0.95, 1.09) | 1.03 (0.97, 1.10) | 1.06 (0.91, 1.23) |
| Latrobe Valley | **1.05 (1.01, 1.09)** | 1.03 (0.99, 1.07) | 1.02 (0.93, 1.12) |
| Background PM$_{2.5}$ | **0.95 (0.93, 0.96)** | **1.01 (1.00, 1.03)** | 1.01 (0.99, 1.04) |
| *Mental health conditions* | | | |
| Morwell | **1.50 (1.37, 1.65)** | **1.12 (1.05, 1.19)** | 1.03 (0.89, 1.18) |
| Latrobe Valley | **1.41 (1.34, 1.48)** | 0.98 (0.94, 1.02) | 0.97 (0.87, 1.07) |
| Background PM$_{2.5}$ | **1.06 (1.04, 1.08)** | 0.99 (0.98, 1.01) | 1.02 (1.00, 1.05) |
| *Injuries* | | | |
| Morwell | **1.11 (1.03, 1.19)** | **1.11 (1.07, 1.16)** | 0.90 (0.77, 1.04) |
| Latrobe Valley | **1.17 (1.12, 1.21)** | **1.07 (1.05, 1.09)** | 0.99 (0.90, 1.08) |
| Background PM$_{2.5}$ | **1.01 (1.00, 1.03)** | **1.01 (1.00, 1.02)** | 1.00 (0.97, 1.03) |

##### Table S5. SENSITIVITY ANALYSIS ADDING BACKGROUND PM$_{2.5}$: Changes in hospital admission, emergency presentations, and ambulance attendance in the eight years following the Hazelwood coalmine fire; continuous model (fire-related PM$_{2.5}$ at Statistical Area level 2)

| | Hospital admissions | Emergency presentations | Ambulance attendance |
|---|---|---|---|
| ***Total*** | | | |
| Fire-related PM$_{2.5}$ | **1.06 (1.05, 1.07)** | **1.08 (1.07, 1.10)** | 1.00 (0.98, 1.01) |
| Background PM$_{2.5}$ | **0.99 (0.98, 0.99)** | **0.98 (0.97, 0.99)** | 0.99 (0.98, 1.01) |
| *Cardiovascular conditions* | | | |
| Fire-related PM$_{2.5}$ | **1.08 (1.05, 1.11)** | **1.05 (1.02, 1.08)** | **0.90 (0.85, 0.95)** |
| Background PM$_{2.5}$ | 1.00 (0.99, 1.01) | 1.01 (0.99, 1.02) | 1.00 (0.98, 1.03) |
| *Respiratory conditions* | | | |
| Fire-related PM$_{2.5}$ | 1.02 (0.99, 1.05) | **1.04 (1.01, 1.07)** | 1.02 (0.96, 1.07) |
| Background PM$_{2.5}$ | **0.95 (0.93, 0.96)** | **1.01 (1.00, 1.03)** | 1.01 (0.98, 1.03) |
| *Mental health conditions* | | | |
| Fire-related PM$_{2.5}$ | **1.31 (1.25, 1.37)** | **1.05 (1.01, 1.08)** | 1.00 (0.95, 1.05) |
| Background PM$_{2.5}$ | **1.06 (1.03, 1.08)** | 0.99 (0.98, 1.01) | 1.02 (0.99, 1.05) |
| *Injuries* | | | |
| Fire-related PM$_{2.5}$ | **1.10 (1.06, 1.13)** | **1.08 (1.06, 1.10)** | 0.96 (0.91, 1.01) |
| Background PM$_{2.5}$ | **1.01 (1.00, 1.03)** | **1.01 (1.00, 1.02)** | 1.00 (0.97, 1.02) |

**Table S6. SENSITIVITY ANALYSIS ADDING BACKGROUND $PM_{2.5}$: Changes in hospital admission, emergency presentations, and ambulance attendance in the eight years following the Hazelwood coalmine fire; categorical model (Morwell and rest of Latrobe Valley compared to rest of regional Victoria)**

| | Hospital admissions | Emergency presentations | Ambulance attendance |
|---|---|---|---|
| ***65+ years*** | | | |
| Morwell | **1.08 (1.04, 1.11)** | **1.07 (1.02, 1.11)** | 1.00 (0.93, 1.08) |
| Latrobe Valley | 1.00 (0.99, 1.02) | **1.02 (1.00, 1.04)** | **0.93 (0.89, 0.97)** |
| Background $PM_{2.5}$ | **0.98 (0.97, 0.99)** | **1.01 (1.00, 1.02)** | 1.00 (0.98, 1.01) |
| *Cardiovascular conditions* | | | |
| Morwell | **1.13 (1.05, 1.21)** | 1.00 (0.92, 1.08) | **0.78 (0.65, 0.95)** |
| Latrobe Valley | **1.04 (1.00, 1.09)** | 0.97 (0.93, 1.01) | 0.92 (0.83, 1.04) |
| Background $PM_{2.5}$ | 0.99 (0.98, 1.01) | **1.03 (1.01, 1.05)** | 1.01 (0.98, 1.04) |
| *Respiratory conditions* | | | |
| Morwell | **1.11 (1.01, 1.23)** | 0.98 (0.89, 1.07) | 1.15 (0.95, 1.39) |
| Latrobe Valley | 0.96 (0.91, 1.02) | 0.86 (0.81, 0.90) | 1.07 (0.95, 1.20) |
| Background $PM_{2.5}$ | **0.95 (0.93, 0.97)** | **1.03 (1.01, 1.05)** | 1.02 (0.98, 1.05) |
| *Mental health conditions* | | | |
| Morwell | **1.60 (1.32, 1.94)** | 1.10 (0.96, 1.26) | 1.32 (0.91, 1.92) |
| Latrobe Valley | **1.46 (1.32, 1.62)** | 1.04 (0.96, 1.12) | 1.00 (0.82, 1.23) |
| Background $PM_{2.5}$ | **1.09 (1.05, 1.13)** | 1.00 (0.97, 1.03) | **1.08 (1.02, 1.16)** |
| *Injuries* | | | |
| Morwell | 0.98 (0.87, 1.10) | **1.12 (1.03, 1.22)** | 0.80 (0.63, 1.00) |
| Latrobe Valley | 1.01 (0.94, 1.07) | **1.13 (1.08, 1.18)** | **0.82 (0.72, 0.93)** |
| Background $PM_{2.5}$ | 0.97 (0.95, 1.00) | **1.04 (1.02, 1.06)** | 0.99 (0.96, 1.03) |
| ***<65 years*** | | | |
| Morwell | **1.11 (1.09, 1.15)** | **1.12 (1.09, 1.16)** | 1.04 (0.97, 1.11) |
| Latrobe Valley | **1.10 (1.09, 1.12)** | **1.12 (1.11, 1.14)** | 0.96 (0.92, 1.01) |
| Background $PM_{2.5}$ | **0.99 (0.98, 0.99)** | **0.98 (0.97, 0.99)** | 0.99 (0.98, 1.00) |
| *Cardiovascular conditions* | | | |
| Morwell | **1.16 (1.07, 1.27)** | **1.15 (1.05, 1.26)** | 0.80 (0.63, 1.03) |
| Latrobe Valley | **1.04 (1.00, 1.09)** | **1.06 (1.01, 1.11)** | 0.89 (0.77, 1.03) |
| Background $PM_{2.5}$ | **1.02 (1.00, 1.04)** | 1.01 (0.99, 1.03) | 1.01 (0.97, 1.05) |
| *Respiratory conditions* | | | |
| Morwell | 0.98 (0.91, 1.07) | 1.02 (0.95, 1.10) | 1.03 (0.83, 1.29) |
| Latrobe Valley | **1.09 (1.04, 1.13)** | **1.11 (1.07, 1.16)** | 0.94 (0.82, 1.08) |
| Background $PM_{2.5}$ | **0.93 (0.92, 0.95)** | **1.02 (1.00, 1.04)** | 1.02 (0.98, 1.07) |
| *Mental health conditions* | | | |
| Morwell | **1.45 (1.31, 1.61)** | **1.09 (1.01, 1.16)** | 1.02 (0.87, 1.19) |
| Latrobe Valley | **1.42 (1.34, 1.50)** | 1.00 (0.96, 1.05) | 0.96 (0.86, 1.07) |
| Background $PM_{2.5}$ | **1.05 (1.03, 1.08)** | 1.01 (0.99, 1.03) | 1.02 (0.99, 1.06) |
| *Injuries* | | | |
| Morwell | **1.19 (1.09, 1.30)** | **1.08 (1.03, 1.13)** | 0.99 (0.82, 1.19) |
| Latrobe Valley | **1.26 (1.21, 1.32)** | **1.09 (1.07, 1.12)** | 1.07 (0.95, 1.20) |
| Background $PM_{2.5}$ | **1.04 (1.02, 1.06)** | **1.01 (1.00, 1.02)** | 1.01 (0.98, 1.04) |

**Table S7. SENSITIVITY ANALYSIS ADDING BACKGROUND $PM_{2.5}$: Changes in hospital admission, emergency presentations, and ambulance attendance in the eight years following the Hazelwood coalmine fire: by age group (65+, <65 years); continuous model (fire-related $PM_{2.5}$ at Statistical Area level 2)**

| | Hospital admissions | Emergency presentations | Ambulance attendance |
|---|---|---|---|
| ***65+ years*** | | | |
| Fire-related $PM_{2.5}$ | **1.04 (1.02, 1.05)** | **1.04 (1.02, 1.06)** | 0.99 (0.96, 1.01) |
| Background $PM_{2.5}$ | **0.98 (0.97, 0.99)** | **1.01 (1.00, 1.02)** | 1.00 (0.99, 1.01) |
| *Cardiovascular conditions* | | | |
| Fire-related $PM_{2.5}$ | **1.08 (1.04, 1.12)** | 1.01 (0.97, 1.05) | **0.91 (0.85, 0.97)** |
| Background $PM_{2.5}$ | 0.99 (0.98, 1.01) | **1.03 (1.01, 1.05)** | 1.01 (0.98, 1.05) |
| *Respiratory conditions* | | | |
| Fire-related $PM_{2.5}$ | 1.04 (0.99, 1.09) | 0.97 (0.93, 1.01) | 1.05 (0.98, 1.12) |
| Background $PM_{2.5}$ | **0.95 (0.93, 0.97)** | **1.03 (1.01, 1.05)** | 1.01 (0.98, 1.04) |
| *Mental health conditions* | | | |
| Fire-related $PM_{2.5}$ | **1.38 (1.26, 1.52)** | **1.07 (1.01, 1.14)** | 1.09 (0.96, 1.24) |
| Background $PM_{2.5}$ | **1.09 (1.05, 1.13)** | 1.00 (0.97, 1.03) | 1.06 (0.99, 1.14) |
| *Injuries* | | | |
| Fire-related $PM_{2.5}$ | 0.98 (0.93, 1.03) | **1.10 (1.06, 1.14)** | **0.91 (0.84, 0.98)** |
| Background $PM_{2.5}$ | 0.97 (0.95, 1.00) | **1.04 (1.02, 1.06)** | 0.98 (0.95, 1.02) |
| **<65 years** | | | |
| Fire-related $PM_{2.5}$ | **1.08 (1.07, 1.10)** | **1.09 (1.08, 1.11)** | 1.01 (0.98, 1.03) |
| Background $PM_{2.5}$ | **0.99 (0.98, 0.99)** | **0.98 (0.97, 0.99)** | 0.99 (0.98, 1.00) |
| *Cardiovascular conditions* | | | |
| Fire-related $PM_{2.5}$ | **1.09 (1.04, 1.13)** | **1.10 (1.06, 1.15)** | 0.92 (0.84, 1.00) |
| Background $PM_{2.5}$ | **1.02 (1.00, 1.04)** | 1.01 (0.99, 1.03) | 1.01 (0.96, 1.05) |
| *Respiratory conditions* | | | |
| Fire-related $PM_{2.5}$ | 1.02 (0.98, 1.06) | **1.06 (1.03, 1.10)** | 1.00 (0.93, 1.08) |
| Background $PM_{2.5}$ | **0.93 (0.92, 0.95)** | **1.02 (1.00, 1.04)** | 1.02 (0.98, 1.07) |
| *Mental health conditions* | | | |
| Fire-related $PM_{2.5}$ | **1.29 (1.23, 1.36)** | **1.04 (1.00, 1.07)** | 0.99 (0.94, 1.05) |
| Background $PM_{2.5}$ | **1.05 (1.03, 1.08)** | 1.01 (0.99, 1.03) | 1.02 (0.99, 1.05) |
| *Injuries* | | | |
| Fire-related $PM_{2.5}$ | **1.17 (1.12, 1.22)** | **1.07 (1.05, 1.09)** | 0.99 (0.93, 1.06) |
| Background $PM_{2.5}$ | **1.04 (1.02, 1.06)** | **1.01 (1.00, 1.02)** | 1.01 (0.98, 1.05) |

**Table S8. SENSITIVITY ANALYSIS ADDING BACKGROUND $PM_{2.5}$: Changes in hospital admission, emergency presentations, and ambulance attendance in the eight years following the Hazelwood coalmine fire: by sex (women, men); categorical model (Morwell and rest of Latrobe Valley compared to rest of regional Victoria)**

| | Hospital admissions | Emergency presentations | Ambulance attendance |
|---|---|---|---|
| ***Women*** | | | |
| Morwell | **1.08 (1.06, 1.12)** | **1.12 (1.09, 1.16)** | 1.04 (0.97, 1.12) |
| Latrobe Valley | **1.09 (1.08, 1.11)** | **1.12 (1.10, 1.14)** | 0.98 (0.94, 1.02) |
| Background $PM_{2.5}$ | **0.98 (0.98, 0.99)** | 0.99 (0.98, 1.00) | 1.00 (0.98, 1.01) |
| *Cardiovascular conditions* | | | |
| Morwell | **1.15 (1.06, 1.25)** | 1.00 (0.92, 1.09) | 0.81 (0.66, 1.01) |
| Latrobe Valley | **1.10 (1.05, 1.15)** | **1.07 (1.02, 1.12)** | 0.96 (0.84, 1.09) |
| Background $PM_{2.5}$ | **1.02 (1.00, 1.04)** | **1.03 (1.01, 1.05)** | 1.01 (0.98, 1.05) |
| *Respiratory conditions* | | | |
| Morwell | 1.02 (0.94, 1.12) | 1.08 (1.00, 1.16) | 1.20 (0.98, 1.48) |
| Latrobe Valley | **1.05 (1.00, 1.10)** | **1.08 (1.03, 1.13)** | 1.05 (0.92, 1.19) |
| Background $PM_{2.5}$ | **0.93 (0.91, 0.95)** | **1.02 (1.00, 1.04)** | 1.01 (0.98, 1.05) |
| *Mental health conditions* | | | |
| Morwell | **1.75 (1.55, 1.99)** | **1.10 (1.01, 1.20)** | 1.21 (1.00, 1.46) |
| Latrobe Valley | **1.66 (1.55, 1.78)** | 1.04 (0.99, 1.09) | 1.06 (0.94, 1.21) |
| Background $PM_{2.5}$ | **1.07 (1.04, 1.10)** | 1.01 (0.99, 1.03) | **1.05 (1.01, 1.09)** |
| *Injuries* | | | |
| Morwell | 1.00 (0.90, 1.11) | **1.06 (1.01, 1.12)** | 0.93 (0.75, 1.14) |
| Latrobe Valley | **1.12 (1.06, 1.19)** | **1.09 (1.06, 1.12)** | 0.96 (0.85, 1.09) |
| Background $PM_{2.5}$ | 0.99 (0.97, 1.02) | **1.02 (1.01, 1.03)** | 1.01 (0.98, 1.05) |
| ***Men*** | | | |
| Morwell | **1.08 (1.04, 1.11)** | **1.09 (1.05, 1.12)** | 0.97 (0.90, 1.04) |
| Latrobe Valley | **1.06 (1.04, 1.07)** | **1.10 (1.08, 1.12)** | 0.97 (0.93, 1.02) |
| Background $PM_{2.5}$ | 1.00 (0.99, 1.00) | **0.99 (0.98, 0.99)** | 0.99 (0.98, 1.00) |
| *Cardiovascular conditions* | | | |
| Morwell | 1.06 (0.98, 1.15) | 1.08 (0.99, 1.18) | **0.73 (0.59, 0.91)** |
| Latrobe Valley | **1.04 (1.00, 1.08)** | 1.01 (0.97, 1.06) | 0.89 (0.79, 1.01) |
| Background $PM_{2.5}$ | 1.00 (0.98, 1.01) | **1.03 (1.01, 1.05)** | 1.00 (0.97, 1.03) |
| *Respiratory conditions* | | | |
| Morwell | 1.02 (0.94, 1.11) | 0.96 (0.89, 1.03) | 0.98 (0.80, 1.19) |
| Latrobe Valley | 1.04 (0.99, 1.09) | 1.04 (0.99, 1.08) | 1.00 (0.89, 1.13) |
| Background $PM_{2.5}$ | **0.97 (0.95, 0.98)** | **1.03 (1.01, 1.05)** | 1.03 (1.00, 1.07) |
| *Mental health conditions* | | | |
| Morwell | **1.19 (1.06, 1.35)** | 1.07 (0.98, 1.17) | 0.89 (0.72, 1.09) |
| Latrobe Valley | **1.16 (1.08, 1.25)** | 1.00 (0.94, 1.05) | 0.87 (0.75, 1.01) |
| Background $PM_{2.5}$ | **1.06 (1.03, 1.09)** | 1.01 (0.99, 1.03) | 1.02 (0.98, 1.06) |
| *Injuries* | | | |
| Morwell | **1.17 (1.07, 1.28)** | **1.10 (1.05, 1.15)** | 0.89 (0.73, 1.09) |
| Latrobe Valley | **1.22 (1.16, 1.28)** | **1.10 (1.08, 1.13)** | 1.01 (0.90, 1.14) |
| Background $PM_{2.5}$ | **1.03 (1.01, 1.05)** | **1.02 (1.00, 1.03)** | 0.99 (0.96, 1.02) |

#### Table S9. SENSITIVITY ANALYSIS ADDING BACKGROUND $PM_{2.5}$: Changes in hospital admission, emergency presentations, and ambulance attendance in the eight years following the Hazelwood coalmine fire: by sex (women, men) ; continuous model (fire-related $PM_{2.5}$ at Statistical Area level 2)

| | Hospital admissions | Emergency presentations | Ambulance attendance |
|---|---|---|---|
| ***Women*** | | | |
| Fire-related $PM_{2.5}$ | **1.06 (1.05, 1.08)** | **1.09 (1.08, 1.11)** | 1.01 (0.99, 1.03) |
| Background $PM_{2.5}$ | **0.98 (0.98, 0.99)** | 0.99 (0.98, 1.00) | 1.00 (0.98, 1.01) |
| *Cardiovascular conditions* | | | |
| Fire-related $PM_{2.5}$ | **1.10 (1.06, 1.15)** | **1.04 (1.00, 1.08)** | **0.93 (0.86, 1.00)** |
| Background $PM_{2.5}$ | **1.02 (1.00, 1.04)** | **1.03 (1.01, 1.05)** | 1.01 (0.98, 1.05) |
| *Respiratory conditions* | | | |
| Fire-related $PM_{2.5}$ | 1.02 (0.98, 1.07) | **1.08 (1.04, 1.12)** | 1.06 (0.99, 1.15) |
| Background $PM_{2.5}$ | **0.93 (0.91, 0.95)** | **1.02 (1.00, 1.04)** | 1.00 (0.97, 1.04) |
| *Mental health conditions* | | | |
| Fire-related $PM_{2.5}$ | **1.48 (1.40, 1.58)** | **1.05 (1.01, 1.10)** | 1.07 (1.00, 1.14) |
| Background $PM_{2.5}$ | **1.07 (1.04, 1.10)** | 1.01 (0.99, 1.03) | **1.05 (1.01, 1.09)** |
| *Injuries* | | | |
| Fire-related $PM_{2.5}$ | 1.04 (0.99, 1.09) | **1.07 (1.04, 1.09)** | 0.96 (0.90, 1.04) |
| Background $PM_{2.5}$ | 0.99 (0.97, 1.02) | **1.02 (1.00, 1.03)** | 1.01 (0.97, 1.05) |
| ***Men*** | | | |
| Fire-related $PM_{2.5}$ | **1.06 (1.04, 1.07)** | **1.07 (1.05, 1.09)** | 0.98 (0.96, 1.01) |
| Background $PM_{2.5}$ | 1.00 (0.99, 1.00) | 0.99 (0.98, 0.99) | 0.99 (0.98, 1.01) |
| *Cardiovascular conditions* | | | |
| Fire-related $PM_{2.5}$ | **1.06 (1.02, 1.09)** | **1.05 (1.01, 1.10)** | **0.89 (0.82, 0.95)** |
| Background $PM_{2.5}$ | 1.00 (0.98, 1.01) | **1.03 (1.01, 1.05)** | 1.00 (0.97, 1.04) |
| *Respiratory conditions* | | | |
| Fire-related $PM_{2.5}$ | 1.02 (0.98, 1.06) | 1.01 (0.97, 1.04) | 0.99 (0.92, 1.06) |
| Background $PM_{2.5}$ | **0.97 (0.95, 0.98)** | **1.03 (1.01, 1.05)** | 1.03 (1.00, 1.07) |
| *Mental health conditions* | | | |
| Fire-related $PM_{2.5}$ | **1.13 (1.06, 1.20)** | **1.04 (1.00, 1.08)** | 0.94 (0.88, 1.01) |
| Background $PM_{2.5}$ | **1.06 (1.03, 1.09)** | 1.01 (0.99, 1.03) | 1.01 (0.97, 1.06) |
| *Injuries* | | | |
| Fire-related $PM_{2.5}$ | **1.14 (1.10, 1.19)** | **1.08 (1.06, 1.11)** | 0.96 (0.89, 1.03) |
| Background $PM_{2.5}$ | **1.03 (1.01, 1.05)** | **1.02 (1.00, 1.03)** | 0.99 (0.95, 1.02) |

#### 4.1 Socioeconomic variance in the Latrobe Valley

The map in Figure S10 illustrates the distribution in socioeconomic deprivation between Morwell and the rest of the Latrobe Valley based on Index of Relative Socio-economic Advantage and Disadvantage (IRSAD) in 2021 (5). Morwell is in the 2nd percentile for Victoria, indicating it is one of the most deprived areas in the state. The next lowest in the Latrobe Valley is Moe/Newborough, which is in the 5th percentile. The remaining areas range between the 21st and 42nd percentiles (Churchill: 27th, Traralgon [East]: 21st, Traralgon [West]: 42nd, Yallourn North/Glengarry: 30th).

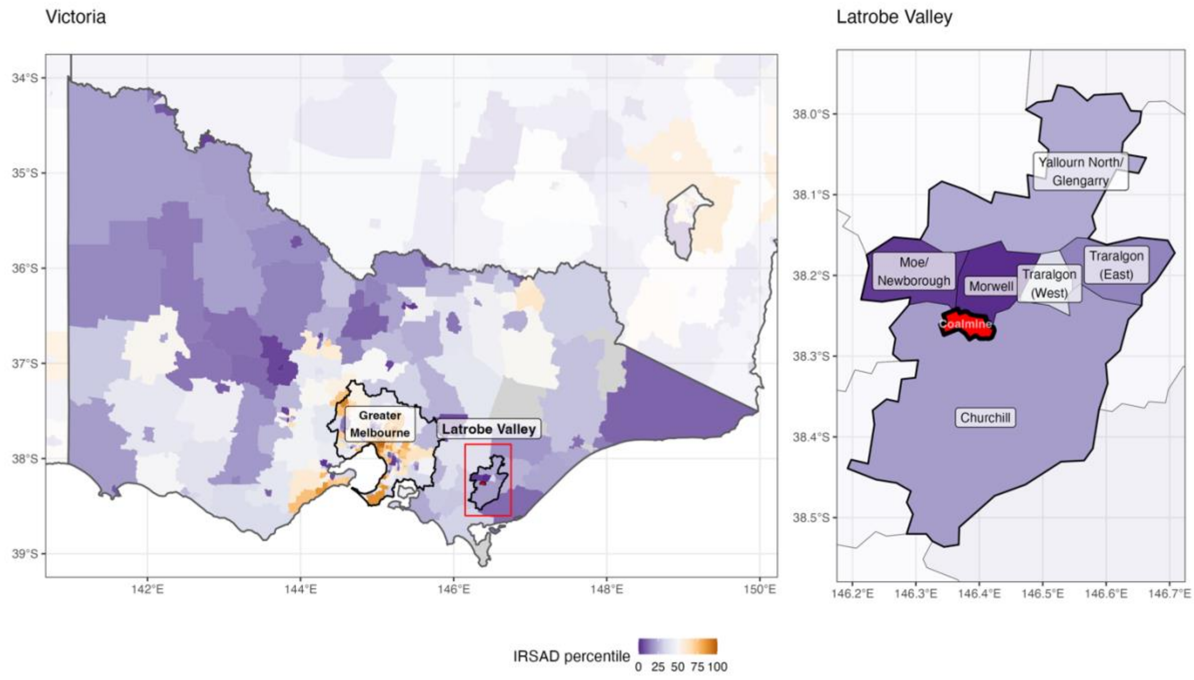

**Figure S10. Map of Victorian socioeconomic (IRSAD) rankings in the Latrobe Valley**

#### 4.3 Background $PM_{2.5}$ often associated with reduced service use

In sensitivity analyses that added background $PM_{2.5}$ to regression models, we found it was frequently associated with *reduced* service use. This is in contrast with an extensive body of literature linking $PM_{2.5}$ to poorer health outcomes and health service use (6,7). We suspected the effect may have been due to complex associations between air pollution and socioeconomic status. For instance, Figure S3 presents the correlation between Index of Relative Socioeconomic Advantage and Disadvantage (IRSAD) (5) and background $PM_{2.5}$ (3) at Statistical Area Level 2. Both datasets were from 2021 to account for events that may have affected Morwell's socioeconomic standing, namely the closure of the coalmine and power plant in 2017. These analyses are presented in Figure S3.

When taken as a whole, more advantaged areas in Victoria have higher levels of background $PM_{2.5}$, likely due to greater amounts vehicle traffic and industrial activity; a standard deviation increase in the score (indicating greater advantage) was associated with an 8.3% (95%CI: 7.0% to 9.4%) increase in background $PM_{2.5}$; this is reflected by the grey line in Figure S3. However, there were clear differences between Greater Melbourne (blue dots), in which background $PM_{2.5}$ levels were generally higher and indicative of a positive association with socioeconomic status and regional Victoria (orange dots and line), where there was no detectable association (-0.9%, 95%CI: -2.1% to 0.3%).

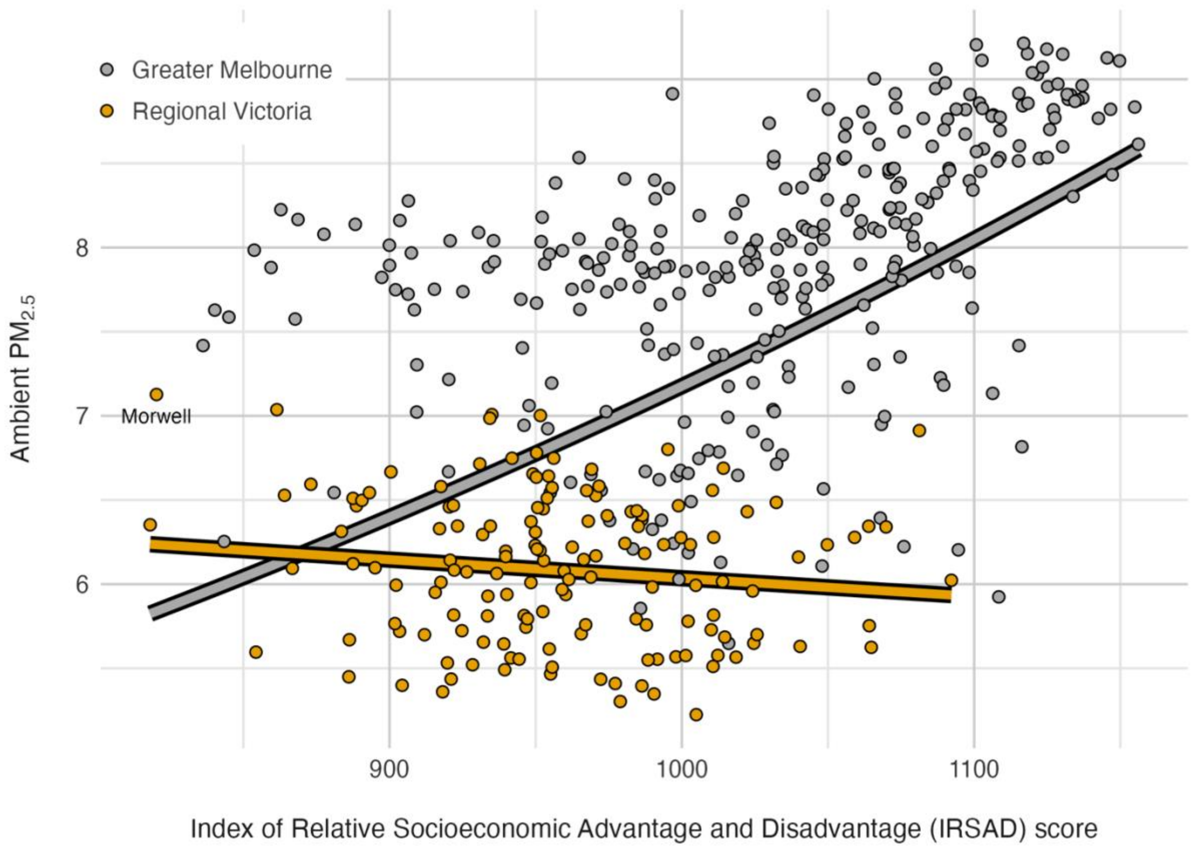

**Figure S11. Relationship between socioeconomic status (based on IRSAD) and ambient $PM_{2.5}$**

Another potential explanation for the unexpected negative associations between background $PM_{2.5}$ and health service use is aggregation error. In other words, the overall level of background $PM_{2.5}$ across Statistical Areas may poorly reflect actual exposures of people in a given area, since they ignore heterogeneity, particularly between populated areas with more vehicular/industrial activity and less populated areas. This problem is exaggerated in regional areas, since Statistical Areas are population-defined, using sizes that are designed to be roughly comparable, ranging from 3,000 to 25,000 people but averaging around 10,000 (8). This means they are geographically larger than urban Statistical Areas and background $PM_{2.5}$ estimates at this level are less accurate. Further, as they have considerably more unpopulated space, their estimates are further biased downwards.
